# Global burden of stigma and discrimination against transgender and gender-diverse adults: a systematic review and meta-analysis

**DOI:** 10.64898/2026.04.22.26351490

**Authors:** Maclaine Barré-Quick, Ping Teresa Yeh, Caitlin E. Kennedy, Hiroshi Azuma, Cortney McLellan, Erin E. Cooney

## Abstract

**Importance:** Stigma and discrimination against transgender and gender-diverse people are prevalent across many settings and may contribute to substantial health disparities.

**Objective:** To synthesize global evidence on the prevalence of stigma, discrimination, and resilience among transgender (trans) and gender-diverse adults.

**Data Sources:** A systematic search was conducted in PubMed, Embase, CINAHL, Cochrane Central, LILACS, and PsycInfo for articles published between January 1, 2010 and January 2, 2023. This database search was supplemented by grey literature and secondary reference searches.

**Article Selection:** Studies were eligible if they presented primary quantitative data on prevalence of stigma, discrimination, and/or resilience among trans and gender-diverse adults (aged 18 and over), with no restrictions on study design, language, or geographic region.

**Data Extraction and Synthesis:** Two independent reviewers extracted data using standardized forms, with discrepancies resolved by consensus. The JBI Critical Appraisal Checklist for Prevalence Articles was used to assess risk of bias. Random effects meta-analysis was conducted for dichotomous prevalence measures using inverse variance weighting and logit transformation; non-dichotomous prevalence data were summarized descriptively.

**Main Outcomes and Measures:** Outcomes included prevalence estimates for various forms of stigma (anticipated, perceived, internalized, and experienced), discrimination in legal/institutional settings (housing, healthcare, employment, police/prison), and resilience.

**Results:** A total of 97 articles, with data from 72,158 unique trans and gender-diverse participants across 26 countries, met inclusion criteria. Studies showed moderate levels of anticipated stigma, perceived stigma, and internalized stigma. Meta-analyses of 36 studies provided pooled estimates of discrimination prevalence across multiple domains: 21.4% in housing (e.g., eviction, rental denial), 24.6% in healthcare (e.g., denial of care, mistreatment), 32.8% in employment (e.g., hiring bias, workplace harassment), and 39.1% in police/prison settings (e.g., profiling, mistreatment). High heterogeneity was observed across studies, reflecting regional and methodological differences. Resilience scores ranged from moderate to high, indicating variation within trans and gender-diverse communities.

**Conclusions and Relevance:** This systematic review and meta-analysis found that stigma and discrimination against trans and gender-diverse adults are pervasive globally. Variation in stigma and discrimination across settings and regions underscores the need for targeted interventions and policy reforms.

**Funding:** World Health Organization through a grant from the Elton John AIDS Foundation and the Bill and Melinda Gates Foundation.

**Key Points:** Question: What is the global prevalence of stigma, discrimination, and resilience among trans and gender-diverse adults?

Findings: A systematic review of 97 articles, and meta-analysis of a subset of 36 articles, found elevated prevalence of stigma in non-institutional settings, discrimination in institutional settings, and resilience.

Meaning: These findings illustrate that stigma and discrimination against trans and gender-diverse adults are pervasive globally and should be addressed through targeted interventions and policy reform.

## Introduction

Stigma and discrimination against trans and gender-diverse adults are significant global public health issues perpetuating health inequities. Systemic barriers, including societal stigma, cultural biases, and discriminatory practices, restrict access to health care, employment, housing, and social resources.^1^ Lack of legal protections in many settings, coupled with punitive policies and inadequate societal support, often exacerbate these disparities, leaving many trans and gender-diverse individuals marginalized and without essential resources for well-being.^1-3^ Discrimination within healthcare systems can lead to care avoidance or delays, as fear of mistreatment and lack of affirming care contribute to untreated health conditions, reinforcing health inequities. ^3-6^

While prior systematic reviews have examined aspects of stigma and discrimination against trans and gender-diverse people, such as the impact of bias and prejudice on mental health outcomes^7^ and discrimination within healthcare systems,^8^ there remains a critical gap in synthesizing the burden of stigma and discrimination globally. To our knowledge, no systematic review has comprehensively synthesized these data across diverse geographic and sociopolitical contexts. Addressing this gap is essential for understanding the burden of stigma and discrimination, informing policy, and designing effective interventions. Here, we assess data on the global prevalence of stigma, discrimination, and resilience among trans and gender-diverse adults.

## Methods

This systematic review and meta-analysis of the prevalence of stigma, discrimination, and resilience among trans and gender-diverse individuals is one component of a broader evidence review on the prevalence and health impact of interpersonal violence, stigma, and discrimination experienced by trans and gender diverse adults globally. We followed PRISMA 2020 guidelines for conducting and reporting systematic reviews and meta-analyses.^9^

### Definitions

We adopted UNAIDS’s definition of stigma as a “dynamic process of devaluation that significantly discredits an individual in the eyes of others,”^10^ and defined discrimination as the enactment of this process. Six categories of stigma, discrimination, and resilience were adapted from the STRIVE model:^11^ (1) anticipated stigma, (2) perceived stigma, (3) internalized stigma, (4) experienced stigma outside legal/institutional settings, (5) discrimination within legal/institutional settings, and (6) resilience.

### Eligibility Criteria

We included primary studies of any design and from any geographic region that presented quantitative prevalence data on stigma, discrimination, and/or resilience among trans and gender-diverse adults (aged ≥18 years). We excluded review articles, editorials, commentaries, studies with fewer than 10 trans and gender-diverse participants, and studies that exclusively reported experiences before age 18 or that did not disaggregate adult data.

### Study Selection

We searched PubMed, Embase, CINAHL, Cochrane Central, LILACS, and PsycInfo to identify all relevant publications in any language from Jan 1, 2010 through Jan 2, 2023, searched grey literature and performed secondary searches of article references and relevant systematic reviews (search strategy details in Appendix A).^12-14^

After removing duplicates, a member of the study team performed an initial round of screening of the titles, abstracts, citation information, and descriptor terms for the citations identified by our search to remove articles that were plainly irrelevant (e.g., pre-clinical studies). Two independent reviewers then assessed title/abstracts for eligibility, with consensus, before two independent reviewers assessed the full texts of all potentially eligible articles for final inclusion, with differences resolved through consensus.

### Data Extraction and Quality Assessment

Data were extracted independently from eligible articles by two reviewers using standardized forms in Microsoft Excel. Differences in data extraction were resolved through consensus and, when necessary, referral to a senior study team member. We adapted the JBI Critical Appraisal Checklist for Prevalence Articles to assess risk of bias,^15^ scoring the nine criteria as “Yes”=1 and “Not applicable”, “Not reported”, or “No”=0.

### Population

We defined trans and gender-diverse individuals as those whose gender identity differs from their sex assigned at birth. For harmonization across studies, *trans women* included those identifying as women, trans women, or other binary feminine-spectrum identities assigned male at birth; *trans men* included those identifying as men, trans men, or masculine-spectrum identities assigned female at birth; and *non-binary* individuals were those not identifying with a singular binary gender, regardless of sex assigned at birth.

### Statistical Analysis

Due to heterogenous study designs and populations, we used random effects meta-analysis to estimate pooled prevalence. Studies were pooled for analysis using inverse variance weighting and logit transformation of proportions based on crude numerators and denominators. A quantile of the standard normal distribution was used to calculate 95% confidence intervals. Between-article variance was estimated using the restricted maximum likelihood method, as this results in less bias in the context of high heterogeneity and large differences in sample size.^16,17^ For articles that reported data from scales, if a standardized cut-point to indicate severity existed, we deferred to these cut-points. For scales without standardized cut-points, we considered scores up to 1/3 of the possible maximum score as low, from 1/3 to 2/3 as moderate, and 2/3 to maximum as high.

We meta-analyzed data from studies with sufficiently similar definitions of each category of stigma, discrimination, and/or resilience as dichotomous prevalence outcomes. When multiple articles reported prevalence estimates drawn from the same study, the estimate reflecting the broadest population and broadest measure was selected for meta-analysis. Statistical analyses were performed using the metaprop function of the *meta* package 8.0-1 in R version 4.3.2.^18,19^

## Results

A total of 72,158 participants from 26 countries were represented in the 97 included articles, comprising 80 unique studies (Figure 1). All studies employed cross-sectional survey designs (Appendix B). The majority (74%, k=73/97) sampled general populations of trans and gender-diverse individuals, followed by studies focused exclusively on trans women (23%, k=23/97) and trans men (2%, k=2/97; Table 1). One-third (28%, k=27/97) of articles reported including non-binary individuals.

**Table 1.**
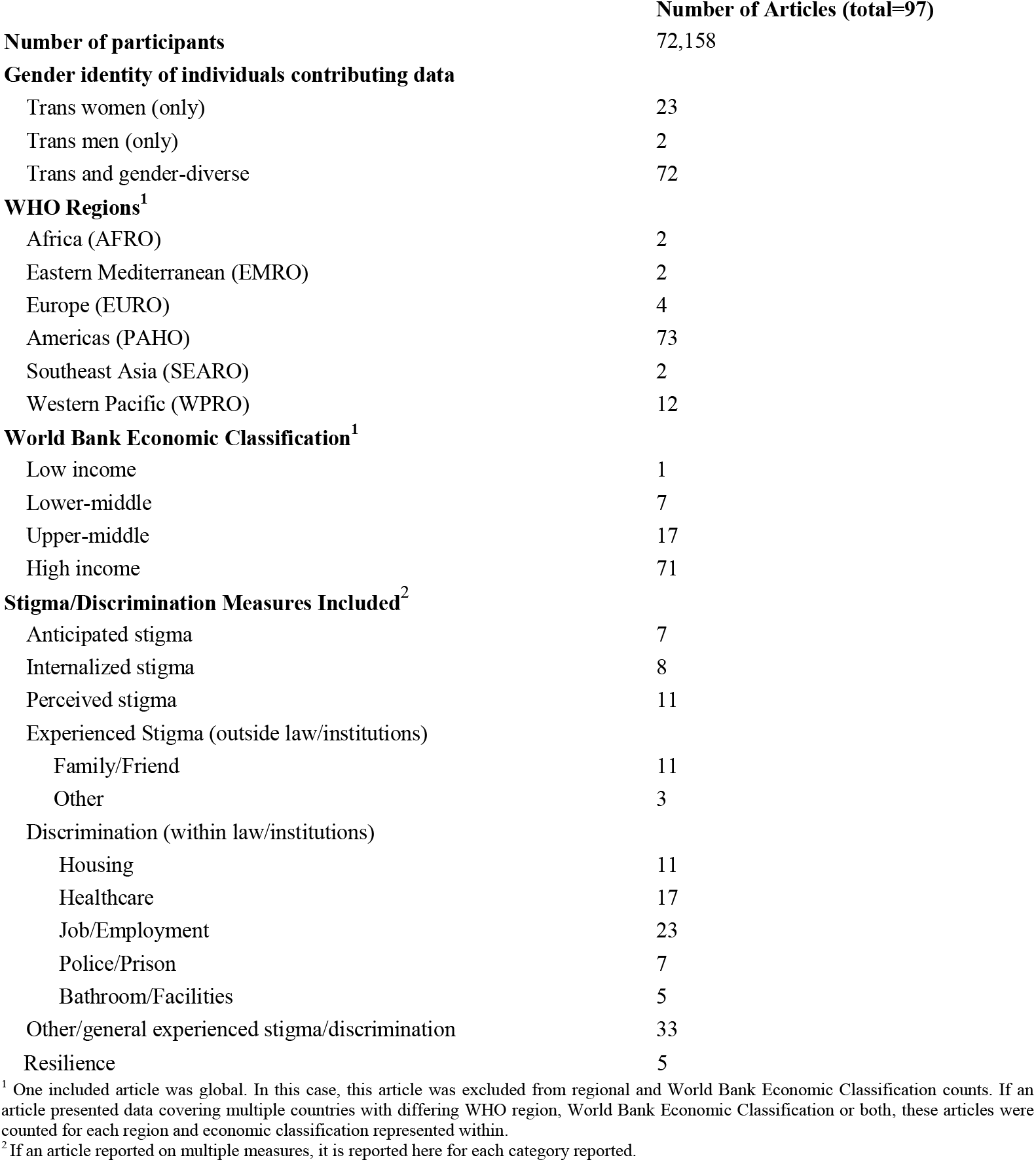
Characteristics of articles included in the review (k=97)

**Figure 1:**
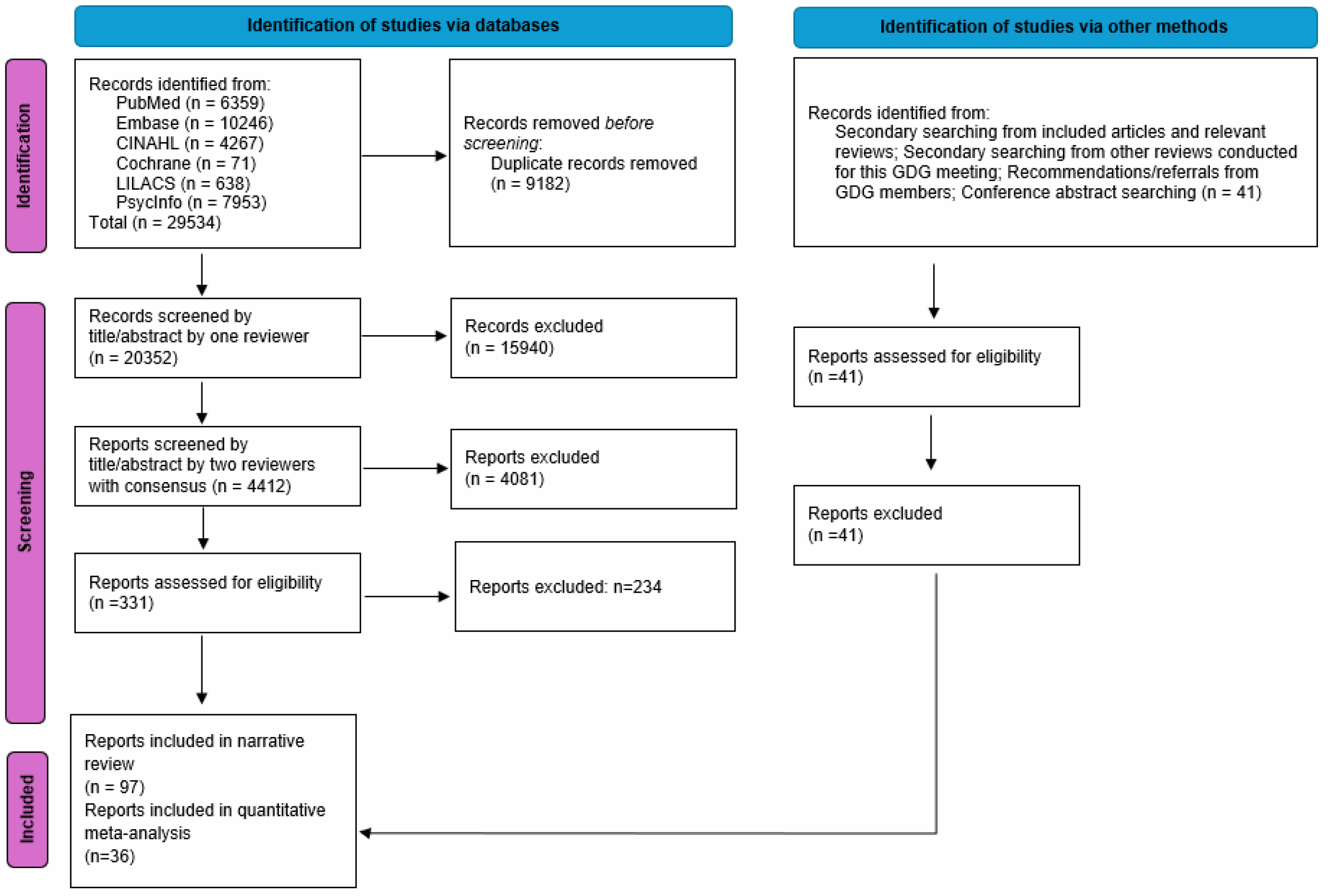
PRISMA diagram showing disposition of citations through the search and screening process.

Most data (74%, k=72/97) were from high-income countries, with nearly two-thirds (64%, k=62/97) from the United States (Figure 2). Fewer articles originated from upper-middle-income (18%, k=17/97) and low- or lower-middle-income settings (8%, k=8/97). Most articles were assessed to have a moderate risk of bias: median risk of bias score was 4 (IQR: 3.5-4). Detailed assessments are presented in Appendix C. Reliance on convenience sampling and small sample sizes limited representativeness and generalizability, which impacted risk of bias scores.

**Figure 2:**
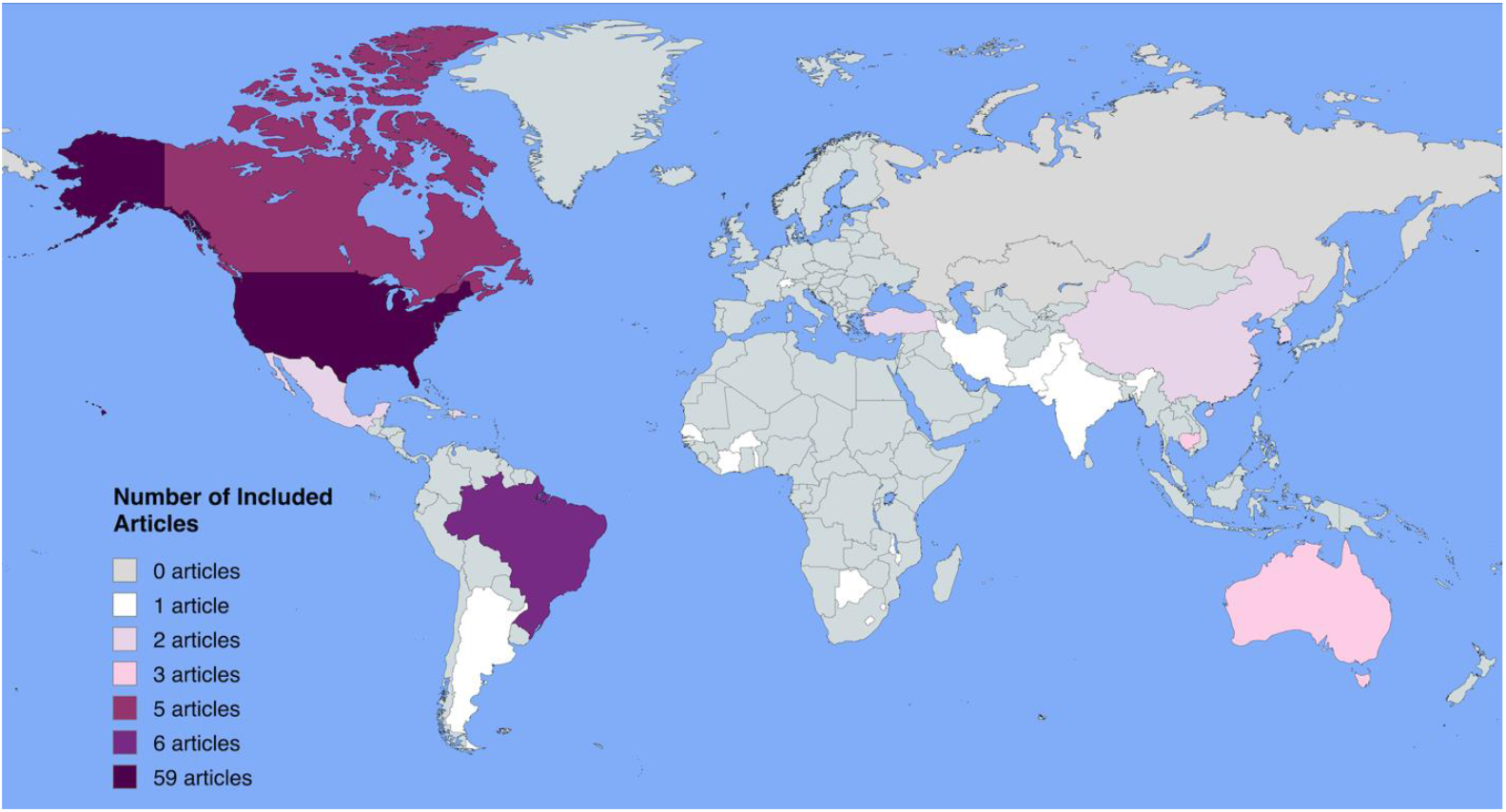
Map of articles published between 2010 and 2023 that reported prevalence of stigma/discrimination/resilience among trans and gender diverse adults.

Stigma and discrimination were assessed using a range of self-reported measures. Nearly two-thirds (59%, k=58/97) reported dichotomous outcomes (e.g., presence or absence of stigma or discrimination), while 38% (k=37/97) reported continuous scores; three articles reported both. Dichotomous prevalence data from 36 articles contributed to meta-analyses; the remaining 22 used outcome definitions that were not comparable.

### Anticipated Stigma

Seven articles measured anticipated stigma,^20-26^ including avoidance of bathrooms, fear of seeking police assistance, and fear of arrest due to transgender identity. In the United States, one article found 61.9% of participants avoided bathrooms due to fear of experiencing problems while using them.^21^ In South Korea, one article found 47.0% of participants reported avoiding bathrooms for fear of experiencing stigma.^27^ In the United States, 26.1% reported fear of seeking police assistance as a transgender person,^23^ while in Cambodia, fear of arrest due to transgender identity was reported by nearly a quarter of participants (24.8%).^26^ In both the United States and China, one article assessed fear of stigma when accessing health services, with findings of 63.3% and 54.8%, respectively.^22,25^ Finally, one article from Mexico found almost half (46.0%) of participants reported fear of being made fun of or called names for being transgender.^24^

### Internalized Stigma

Eight articles measured internalized stigma.^28,77,88,24,29-32^ Three articles, conducted in Argentina, Mexico, and the United States, reported prevalance of internalized stigma ranging from 16.0%-56.0%.^24,29,30^ Five studies reported scale measures of internalized stigma. Four studies, including two from the United States, one from Switzerland and one from Italy, used the Gender Minority Stress and Resilience Scale, specifically the internalized transphobia subscale (range: 0-40). They found moderate levels of internalized stigma in a sample in the United States with a mean score of 16.8 (SD: 6.7), low levels in Switzerland with a mean score of 7.8, and low levels in Italy with a mean score of 12.1 (SD: 8.7).^28,33,34^ One article from the United States used a modified Homosexual Stigma Scale, ranging from 1 to 4, and reported a mean score of 1.8 (SD: 0.7), indicating moderate levels of internalized stigma.^32^ The final article from the United States found a moderate level of internalized stigma with a mean of 4.3 (SD: 3.2), using a modified 2-item version for the Gender Minority Stress and Resilience Scale (range 0-12).^31^

### Perceived Stigma

Eleven articles reported perceived stigma, all measured using scales.^32,35-44^ Four articles, all from the United States, used the Everyday Discrimination Scale, which ranges from 0-44.^35,40,43,44^ Reported mean scores ranged from 12.7 to 19.9, indicating low to moderate levels of perceived stigma. The remaining articles from the United States used unique scales and reported means that indicate moderate levels of perceived stigma.^32,37,38^ In Turkey, one article reported moderate levels of personal stigma (Perceived Personal Discrimination Scale: mean 9.5, SD: 4.0, range 4–20) and greater group-level stigma (Perceived Group Discrimination Scale: mean 25.8, SD: 6.5, range 7–35).^36^ In China, an article using a self-perceived discrimination scale found moderate levels of perceived stigma, with a mean score of 51.4 (SD: 2) on a scale ranging from 0 to 100.^45^ In Brazil, one article reported a moderate discrimination score mean of 3.1 (SD: 2.1, range 0-8).^39^ In Canada, an article using a 10-item (range 0-30) transphobia scale reported a mean score of 13.7 (SD: 6.0), indicating moderate stigma levels.^41^

### Discrimination Within Legal and Institutional Environments

Sixty-seven articles reported discrimination within specific institutional environments, such as housing, healthcare, and employment. Table 2 presents meta-analysis results across these settings (accompanying forest plots in Appendix D).

**Table 2:**
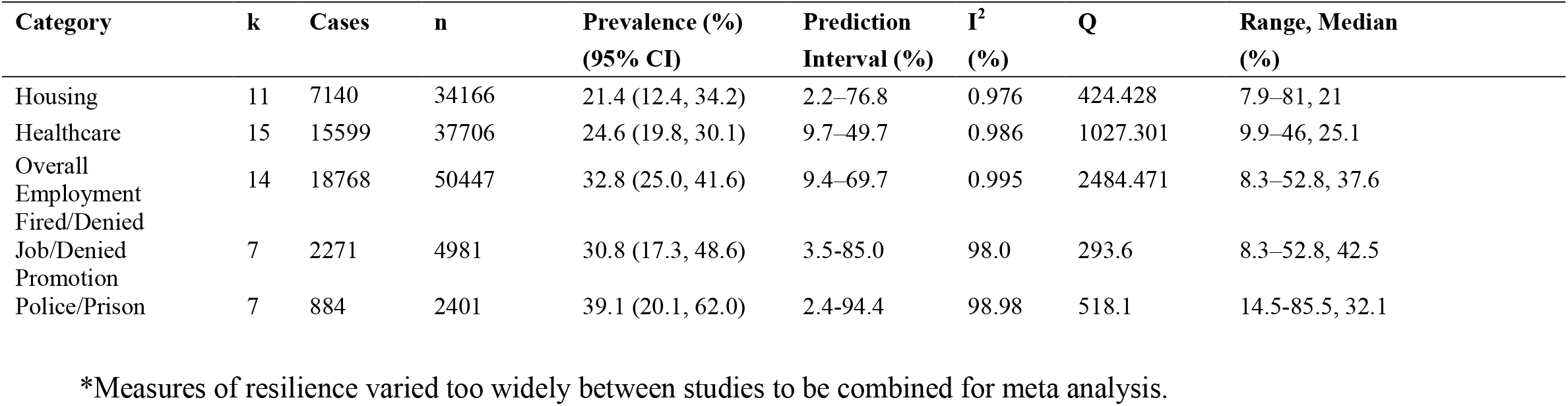
Pooled prevalence of discrimination in trans and gender-diverse adults.

In the housing domain, measures of discrimination included rental denial, eviction, and housing instability due to gender identity. In meta-analysis of data from 11 articles, the pooled prevalence of housing discrimination was 21.4% (95% CI: 12.4–34.2) (Appendix D1).

In the healthcare domain, measures of discrimination included care denial, harassment by healthcare providers, and inadequate treatment due to gender identity. In meta-analysis of data from 17 articles, the pooled prevalence of healthcare discrimination was 24.6% (95% CI: 19.8– 30.1) (Appendix D2).

In the general employment domain, measures of discrimination included workplace harassment, unjust termination, and barriers to hiring. In meta-analysis of data from 14 articles, the pooled prevalence of general employment discrimination was 32.8% (95% CI: 25.0–41.6) (Appendix D3). One article among 78 transgender and gender-diverse individuals in the United States found low work or school-related harassment scores averaging 2.21 (SD = 1.10) on a scale from 1 (the event never happened to you) to 6 (the event happened almost all the time).^46^ In meta-analysis of seven articles specifically measuring denial of job opportunities or promotions, pooled prevalence of denial of job opportunities was 30.8% (95% CI: 17.3–48.6) (Appendix D4).

In interactions with police and the prison system, measures of discrimination included mistreatment, profiling, and lack of access to adequate support due to gender identity. In meta-analysis of pooled data from seven articles, pooled prevalence of discrimination by police or in prisons was 39.1% (95% CI: 20.1–62.0) (Appendix D5).

### Experienced Stigma Outside Legal and Institutional Environments

Forty-two articles reported experienced stigma outside of legal and institutional contexts. An article presenting data from 937 transgender women from seven African countries reported high levels of discrimination within family and friend circles: 35.0% reported experiencing disparaging remarks or gossip from family members, 29.8% felt rejection from friends, and 17.8% had been excluded from family gatherings due to their identities.^47^ The prevalence of family-related stigma was similar in other contexts. In Brazil, 51.6% of a sample of 670 transgender and gender-diverse adults reported experiencing derogatory language from family members.^48^ In China, an article presenting data from of 200 transgender women found 70.5% encountered family-related discrimination, while 30.5% reported friend-related discrimination.^49^ In Latin America, studies revealed similar patterns of discrimination, with nearly half of a sample of 291 transgender women in the Dominican Republic reporting that they had lost close friendships (47.8%) or faced family conflicts (43.3%) due to their gender identity, ^54^ while 33.1% of 148 transgender individuals in Mexico experienced stigma from neighbors and 30.4% were excluded from their families.^50,51^ In a sample of 539 transgender women working as sex workers in the United States, 67.3% had experienced family hurt or embarrassment related to their gender identity.^52^ Another article presenting data from the United States used the Relationship Stigma Scale (range: 0–18) to assess stigma in relationships between transgender women and their male partners and reported a mean score of 6.9 (SD: 6.6).^53^

### Other/General definitions of experienced stigma/discrimination

Thirty-three articles measured a different type of stigma and discrimination or defined stigma or discrimination in a way that was too distinct to combine into another category.^20,27-29,33,54-80^ Eight of these articles, three from the United States, three from Brazil and one from Cambodia, used a general definition of any kind of discrimination over any period of time (e.g. “ever discrimination,”) as a dichotomous outcome, producing prevalences of experienced discrimination between 38.5% and 95.7%; six articles reported prevalence over 85.0%. ^20,54,56,57,61,63,76,79^ Sixteen articles used a variety of scales to assess experienced discrimination (e.g., “lifetime transphobia, “enacted stigma,” “transphobia,” “discrimination experiences,”), presenting moderate values.^28,29,55,62,66,69-71,73-75,77-80^

### Resilience

Three articles reported resilience, all using scales.^51,75,81-83^ An article presenting data from 137 transgender individuals in Jamaica assessed using the Resilient Coping Scale, which ranges from 6 to 30, reported a mean score of 19.90 (SD: 5.0) indicating moderately high resilience.^81^ Similarly, using the Resilience Scale (range: 0–40), a sample of 42 transgender and gender-diverse individuals reported a mean score of 32.2 (SD: 2.0), indicating high levels of resilience.^70^ Finally, a sample of 116 transgender and gender-diverse individuals in Turkey assessed on the Resilience Scale for Adults, which is 33 items, each ranging 1 to 5, produced a mean score of 128.62 (SD: 20.26) indicating high resilience.^82^

## Discussion

In this systematic review and meta-analysis, we found stigma and discrimination were highly prevalent among trans and gender-diverse individuals across various domains. These findings align with previous research documenting the burden of stigma and discrimination.^1,84,85^ Although the pooled prevalences from the meta-analyses cannot be interpreted as global averages, they offer insight into the burden across regions and domains of stigma and discrimination. The high heterogeneity of prevalence estimates and definitions of experienced stigma and discrimination observed across articles is notable. While some countries have enacted policies to reduce discrimination, others impose legal barriers that reinforce societal stigma and impede access to essential resources.^86^

We found high levels of housing discrimination reflecting systemic exclusion from safe and stable housing, which has been linked to increased rates of homelessness and associated health risks.^87^ Legal protections are necessary to protect trans and gender diverse individuals from discriminatory housing policies and practices by landlords.^88,89^ High levels of healthcare discrimination were also reported in various forms: denial of care, harassment by providers, and inadequate treatment. Healthcare discrimination contributes to delays in seeking care, avoidance of medical settings, and poor health outcomes.^90^ Sensitivity training for providers and legal protections are required to create the institutional structure that safeguards against stigma and discrimination against trans and gender diverse people in institutional settings, including healthcare systems.^90,91^ Similarly, most studies found high levels were present in employment. Reports of unjust firing or denial of promotions and hiring barriers suggest the need for stronger workplace protections and enforcement mechanisms to reduce barriers to career advancement and economic security for trans and gender-diverse individuals.^92^ Addressing these challenges would require not only anti-discrimination laws but also robust implementation and enforcement mechanisms to ensure safe and supportive environments.

Anticipated stigma in bathrooms, specifically fear of confrontation or victimization, recurred across articles.^20,21^ Gender-neutral bathroom facilities and protective policies would enable trans and gender-diverse individuals to navigate public spaces without fear of confrontation or denial of access.^93-95^ Additionally, family-and friend-related discrimination was common, which has previously been shown to lead to social isolation and worse mental health outcomes.^48,96^ Efforts to support personal networks and reduce familial rejection are crucial to mitigating the social isolation and mental health challenges associated with stigma.^97^

The main strength of this review is its comprehensive approach, incorporating global data and a synthesis of definitions for stigma, discrimination, and resilience. However, findings must be interpreted in light of several limitations. Measures of stigma and discrimination are generally self-reported and rely on the participant’s perception that a certain experience was the result of their gender identity, which may not be the case in all instances. Conversely, subtle or indirect discrimination may have gone unnoticed. The role of resilience and coping mechanisms emerged as an under-researched area, highlighting the need for studies that quantify how transgender and gender-diverse individuals adapt to and manage stigma.^98^ Future studies employing systematic and random sampling methods, such as those used in the Trans Pop Survey,^99^ could address generalizability and improve the reliability of findings. Additionally, large convenience samples, such as the US Transgender Survey, which feature racial, ethnic, age, and geographic diversity, may more closely resemble the diversity of the population and allow for subgroup analyses.^100^ Finally, most included articles in this review came from high-income countries, particularly the United States; the scarcity of data in other settings limits the generalizability of findings.

The high prevalence of stigma and discrimination experiences across multiple settings highlights a critical need for inclusive policies and supportive practices for transgender and gender-diverse populations. Addressing both interpersonal stigmas and institutional discrimination is essential for improving health outcomes and enhancing social inclusion. Finally, the concentration of research in high-income countries underscores an urgent need to expand research in low-and middle-income settings to inform equitable global interventions.

## Data Availability

All data produced in the present study are available upon reasonable request to the authors

## Authors’ contributions

This work was conceptualized by CEK, PTY, and EEC. The methodology was designed by CEK, PTY and EEC, and the project administered by PTY. Data were curated by MBQ and HA. Formal analysis was conducted by MBQ, including visualization. MBQ wrote the original draft with support from CEK, PTY and EEC. The final draft was reviewed and edited by all authors.

## Acknowledgements

We thank the Johns Hopkins graduate students and affiliates who assisted in the search, screening, and data abstraction/cleaning process: Lamia Ayaz, Feaven Gebrezgi, Erin Go, Maryam Mansoor, Madhumitha Mathivanan, Anjali Mehta, Jessica Mogi, Kathryn Noon, Zoe Pamonag, Alexandra Quintero, Swapna Roopesh Kumar, Deeksha Tomar, Abigail Ulman, Klonese Williams, Brooke Wong; Ashleigh Rich and Tonia Poteat, who assisted with early conceptualization of this project; and the WHO technical team supporting the guideline development process.

