## Supplementary Materials for "Global burden of stigma and discrimination against transgender and gender-diverse adults: a systematic review and meta-analysis"

**Supplementary material**

Appendix A. Search terms and strategy
Appendix B. Included articles
Appendix C. Risk of bias assessment
Appendix D. Forest plots showing prevalence of discrimination across different sectors experienced by trans and gender-diverse adults

### Appendix A. Search terms and strategy

#### Search terms by database

**PubMed**

**Concept 1: TGD population**

("achout"[tw] OR "AFAB"[tw] OR "agender"[tw] OR "akava'ine"[tw] OR "akava ine"[tw] OR "AMAB"[tw] OR "aravanis"[tw] OR "assigned female at birth"[tw] OR "assigned male at birth"[tw] OR "assigned-female-at-birth"[tw] OR "assigned-female-sex"[tw] OR "assigned-male-at-birth"[tw] OR "assigned-male-sex"[tw] OR "bakla"[tw] OR "bantut"[tw] OR "berdache"[tw] OR "bigender"[tw] OR "brotherboys"[tw] OR "cross dress*"[tw] OR "cross-dress*"[tw] OR "dissident gender*"[tw] OR "fa'afafine"[tw] OR "fa afafine"[tw] OR "fakafifine"[tw] OR "female to male gender"[tw] OR "female to male sex"[tw] OR "female-to-male"[tw] OR "FTM"[tw] OR "gender adjustment"[tw] OR "gender affirm*"[tw] OR "gender atypical"[tw] OR "gender binar*"[tw] OR "gender change*"[tw] OR "gender disorder*"[tw] OR "gender divers*"[tw] OR "gender diversity"[tw] OR "gender dysphor*"[tw] OR "Gender Dysphoria"[Mesh] OR "gender identity disorder*"[tw] OR "gender inclus*"[tw] OR "gender incongruen*"[tw] OR "gender minorit*"[tw] OR "gender neutral*"[tw] OR "gender non binar*"[tw] OR "gender non-binar*"[tw] OR "gender nonbinar*"[tw] OR "Gender Non conform*"[tw] OR "gender non-conform*"[tw] OR "Gender Nonconform*"[tw] OR "gender orientat*"[tw] OR "gender re-assign*"[tw] OR "gender reassign*"[tw] OR "gender variant"[tw] OR "gender affirm*"[tw] OR "gender-affirm*"[tw] OR "gender-bend*"[tw] OR "gender-expansive*"[tw] OR "gender-queer*"[tw] OR "gender queer"[tw] OR "genderdiverse"[tw] OR "genderdiversity"[tw] OR "genderqueer"[tw] OR "gender queer*"[tw] OR "Health Services for Transgender Persons"[Mesh] OR "hijra"[tw] OR "irahuhua"[tw] OR "irawhiti" [tw] OR "kathoey" [tw] OR "Kathoy" [tw] OR "katoey"[tw] OR "khanith" [tw] OR "khwaja sira"[tw] OR "kothi"[tw] OR "koti"[tw] OR "leiti"[tw] OR "mah"[tw] OR "mahuvahine"[tw] OR "mak nyah"[tw] OR "maknyah"[tw] OR "male to female gender"[tw] OR "male to female sex"[tw] OR "male-to-female*"[tw] OR "metis"[tw] OR "minority gender"[tw] OR "MTF"[tw] OR "muxe" [tw] OR "nadleehi" [tw] OR "non-binar*"[tw] OR "non-binary"[tw] OR "non-conform*"[tw] OR "nonbinar*"[tw] OR "nonbinary"[tw] OR "Nonbinary Persons"[tw] OR "nonconform*"[tw] OR "paknyah"[tw] OR "palopa"[tw] OR "people of trans experience"[tw] OR "phuying kham phet"[tw] OR "pinapinaaine"[tw] OR "Queer*"[tw] OR "sao praphet song"[tw] OR "sex change*"[tw] OR "sex re-assign*"[tw] OR "sex reassign*"[tw] OR "Sex Reassignment Procedures"[Mesh] OR "sex-affirm*"[tw] OR "Sexual and Gender Minorities"[Mesh] OR "Sexual Dissident"[tw] OR "sexual dysphor*"[tw] OR "sistergirls"[tw] OR "sworn virgins"[tw] OR "tahine"[tw] OR "takataapui"[tw] OR "tangata ira tane"[tw] OR "tangata ira wahine"[tw] OR "TGD"[tw] OR "third gender*"[tw] OR "third spirit"[tw] OR "third spirited"[tw] OR "trans boy"[tw] OR "trans boys"[tw] OR "trans female*"[tw] OR "trans femin*"[tw] OR "Trans gender*"[tw] OR "trans girl*"[tw] OR "trans male*"[tw] OR "trans masculin*"[tw] OR "Trans sexual"[tw] OR "Trans sexuals"[tw] OR "trans-bod*"[tw] OR "trans-boy*"[tw] OR "trans-fem*"[tw] OR "trans-gender*"[tw] OR "trans-girl*"[tw] OR "trans-ident*"[tw] OR "trans-male*"[tw] OR "trans-man"[tw] OR "trans-men"[tw] OR "trans-people"[tw] OR "trans-person"[tw] OR "trans-persons"[tw] OR "trans-selv*"[tw] OR "trans-sex*"[tw] OR "trans-visib*"[tw] OR "trans-wom*"[tw] OR "transbod*"[tw] OR "transboy*"[tw] OR "transfemal*"[tw] OR "transfemin*"[tw] OR "Transgender Person"[tw] OR "Transgender Persons"[Mesh] OR "transgender*"[tw] OR "transgirl*"[tw] OR "transident*"[tw] OR "transmale*"[tw] OR "transman"[tw] OR "transmasculine"[tw] OR "transmasculinity"[tw] OR "transmen"[tw] OR "transpeople"[tw] OR "transperson"[tw] OR "transpersons"[tw] OR "transpinoy"[tw] OR "transpinoy"[tw] OR "transsex*"[tw] OR "Transsexualism"[Mesh] OR "transvesti*"[tw] OR "transvisib*"[tw] OR "transwom*"[tw] OR "travesti*"[tw] OR "travesty"[tw] OR "two Spirit Persons"[tw] OR "vakasalewalewa"[tw] OR "waria"[tw] OR "whakawahine"[tw] OR "xanith"[tw] OR "2 spirit*"[tw] OR "atypical gender"[tw] OR "bi gender*"[tw] OR "biological female*"[tw] OR "biological male*"[tw] OR "biological man"[tw] OR "biological men"[tw] OR "biological wom*"[tw] OR "brother boys"[tw] OR "gender fluid"[tw] OR "gender incongruen*"[tw] OR "gender questioning"[tw] OR "gender variants"[tw] OR "gender variation"[tw] OR "gender variations"[tw] OR "genderexpans*"[tw] OR "genderfluid"[tw] OR "genderquestioning"[tw] OR "intersex*"[tw] OR "machi"[tw] OR "selrata"[tw] OR "sister girls"[tw] OR "trans experienc*"[tw] OR "trans man"[tw] OR "trans visib*"[tw] OR "transpinay*"[tw] OR "Transvestism"[Mesh])

**Concept 2: stigma/discrimination/violence**

(Violence [Mesh] OR Gender-Based Violence [Mesh] OR violence [tiab] OR abuse [tiab] OR rape [tiab] OR forced sex [tiab] OR coerced sex [tiab] OR molestation [tiab] OR psychological harm [tiab] OR maldevelopment [tiab] OR deprivation [tiab] OR offence [tiab] OR neglect [tiab] OR IPV [tiab] OR Social Stigma [Mesh] OR Discrimination, Psychological [Mesh] OR Social Discrimination [Mesh] OR Perceived Discrimination [Mesh] OR Prejudice [Mesh] OR Stereotyping [Mesh] OR stigma [tiab] OR stigmatization [tiab] OR stigmatisation [tiab] OR stigmatized [tiab] OR stigmatised [tiab] OR discriminat*[tiab] OR oppress*[tiab] OR transphobia [tiab] OR transphobic [tiab] OR prejudice [tiab] OR stereotype [tiab] OR stereotyping [tiab]OR stereotyped attitudes [tiab] OR shame [tiab] OR disrespect [tiab] OR coercive control [tiab] OR forced procedures [tiab] OR interpersonal violence [tiab] OR psychological violence [tiab])

**Embase**

**Concept 1: TGD population**

'sexual and gender minority'/exp OR 'transgender'/exp OR 'gender dysphoria'/exp OR 'sex reassignment'/exp OR 'cross-dressing'/exp OR (('achout':ti,ab,kw OR 'AFAB':ti,ab,kw OR 'agender':ti,ab,kw OR 'akavaine':ti,ab,kw OR 'AMAB':ti,ab,kw OR 'aravanis':ti,ab,kw OR 'assigned-female-at-birth':ti,ab,kw OR 'assigned-female-sex':ti,ab,kw OR 'assigned-male-at-birth':ti,ab,kw OR 'assigned-male-sex':ti,ab,kw OR 'bakla':ti,ab,kw OR 'bantut':ti,ab,kw OR 'berdache':ti,ab,kw OR 'bigender':ti,ab,kw OR 'bi-gender':ti,ab,kw OR 'biological-female':ti,ab,kw OR 'biological-male':ti,ab,kw OR 'biological-man':ti,ab,kw OR 'biological-men':ti,ab,kw OR 'biological-woman':ti,ab,kw OR 'biological-women':ti,ab,kw OR 'brotherboys':ti,ab,kw OR 'brother-boys':ti,ab,kw OR 'cross-dress':ti,ab,kw OR 'dissident-gender':ti,ab,kw OR 'faafafine':ti,ab,kw OR 'fakafifine':ti,ab,kw OR 'female-to-male':ti,ab,kw OR 'gender-affirm':ti,ab,kw OR 'gender-atypical':ti,ab,kw OR 'atypical-gender':ti,ab,kw OR 'gender-bend':ti,ab,kw OR 'gender-binary':ti,ab,kw OR 'gender-change':ti,ab,kw OR 'gender-disorder':ti,ab,kw OR 'gender-diverse':ti,ab,kw OR 'genderdiverse':ti,ab,kw OR 'gender-dysphoria':ti,ab,kw OR 'gender-expansive':ti,ab,kw OR 'genderexpansive':ti,ab,kw OR 'genderfluid':ti,ab,kw OR 'gender-fluid':ti,ab,kw OR 'gender-identity-disorder':ti,ab,kw OR 'gender-inclusive':ti,ab,kw OR 'gender-incongruent':ti,ab,kw OR 'gender-minority':ti,ab,kw OR 'gender-neutral':ti,ab,kw OR 'gender-orientation':ti,ab,kw OR 'genderqueer':ti,ab,kw OR 'genderquestioning':ti,ab,kw OR 'gender-questioning':ti,ab,kw OR 'gender-reassign':ti,ab,kw OR 'gender-re-assign':ti,ab,kw OR 'gender-variant':ti,ab,kw OR 'hijra':ti,ab,kw OR 'irahuhua':ti,ab,kw OR 'intersex':ti,ab,kw OR 'irawhiti':ti,ab,kw OR 'kathoey':ti,ab,kw OR 'kathoy':ti,ab,kw OR 'katoey':ti,ab,kw OR 'khanith':ti,ab,kw OR 'khwaja sira':ti,ab,kw OR 'kothi':ti,ab,kw OR 'koti':ti,ab,kw OR 'leiti':ti,ab,kw OR 'machi':ti,ab,kw OR 'mah':ti,ab,kw OR 'mahuvahine':ti,ab,kw OR 'male-to-female':ti,ab,kw OR 'mak-nyah':ti,ab,kw OR 'maknyah':ti,ab,kw OR 'metis':ti,ab,kw OR 'minority-gender':ti,ab,kw OR 'muxe':ti,ab,kw OR 'nadleehi':ti,ab,kw OR 'non-binary':ti,ab,kw OR 'nonbinary':ti,ab,kw OR 'non-conforming':ti,ab,kw OR 'nonconforming':ti,ab,kw OR 'paknyah':ti,ab,kw OR 'palopa':ti,ab,kw OR 'phuying-kham-phet':ti,ab,kw OR 'pinapinaaine':ti,ab,kw OR 'queer':ti,ab,kw OR 'sao-praphet-song':ti,ab,kw OR 'selrata':ti,ab,kw OR 'sex-affirming':ti,ab,kw OR 'sex-change':ti,ab,kw OR 'sex-reassignment':ti,ab,kw OR 'sex-re-assignment':ti,ab,kw OR 'sexual-dissident':ti,ab,kw OR 'sexual-dysphoria':ti,ab,kw OR 'sistergirls':ti,ab,kw OR 'sister-girls':ti,ab,kw OR 'sworn-virgin':ti,ab,kw OR 'tahine':ti,ab,kw OR 'takataapui':ti,ab,kw OR 'tangata-ira-tane':ti,ab,kw OR 'tangata-ira-wahine':ti,ab,kw OR 'third-gender':ti,ab,kw OR 'third-spirit':ti,ab,kw OR 'transbodies':ti,ab,kw OR 'trans-bodies':ti,ab,kw OR 'transboy':ti,ab,kw OR 'trans-boy':ti,ab,kw OR 'trans-experience':ti,ab,kw OR 'transfeminine':ti,ab,kw OR 'trans-feminine':ti,ab,kw OR 'transgender':ti,ab,kw OR 'trans-gender':ti,ab,kw OR 'transgirl':ti,ab,kw OR 'trans-girl':ti,ab,kw OR 'trans-identity':ti,ab,kw OR 'transidentity':ti,ab,kw OR 'transmale':ti,ab,kw OR 'trans-male':ti,ab,kw OR 'transman':ti,ab,kw OR 'trans-man':ti,ab,kw OR 'transmen':ti,ab,kw OR 'trans-men':ti,ab,kw OR 'transmasculine':ti,ab,kw OR 'trans-masculine':ti,ab,kw OR 'trans-people':ti,ab,kw OR 'transpeople':ti,ab,kw OR 'trans-person':ti,ab,kw OR 'trans-persons':ti,ab,kw OR 'transperson':ti,ab,kw OR 'transpersons':ti,ab,kw OR 'transpinay':ti,ab,kw OR 'transpinoy':ti,ab,kw OR 'trans-selves':ti,ab,kw OR 'transsexual':ti,ab,kw

OR 'trans-sexual':ti,ab,kw OR 'transvestite':ti,ab,kw OR 'transvisibility':ti,ab,kw OR 'trans-visibility':ti,ab,kw OR 'transwoman':ti,ab,kw OR 'trans-woman':ti,ab,kw OR 'travesti':ti,ab,kw OR 'two-spirit':ti,ab,kw OR '2-spirit':ti,ab,kw OR 'vakasalewalewa':ti,ab,kw OR 'waria':ti,ab,kw OR 'whakawahine':ti,ab,kw OR 'xanith':ti,ab,kw) NOT ('male-to-female ratio':ti,ab,kw OR 'male-to-female ratios':ti,ab,kw OR 'female-to-male ratio':ti,ab,kw OR 'female-to-male ratios':ti,ab,kw OR 'transfemoral':ti,ab,kw))

**Concept 2: stigma/discrimination/violence**

'violence'/exp OR 'domestic violence'/exp OR 'dating violence'/exp OR 'emotional abuse'/exp OR 'exposure to violence'/exp OR 'gender based violence'/exp OR 'partner violence'/exp OR 'stigma'/exp OR 'social stigma'/exp OR 'discrimination against sexual and gender minorities'/exp OR 'transphobia'/exp OR 'social discrimination'/exp OR 'perceived discrimination'/exp OR 'prejudice'/exp OR 'stereotyping'/exp OR 'violence':ti,ab,kw OR 'gender-based violence':ti,ab,kw OR 'abuse':ti,ab,kw OR 'rape':ti,ab,kw OR 'forced sex':ti,ab,kw OR 'coerced sex':ti,ab,kw OR 'molestation':ti,ab,kw OR 'psychological harm':ti,ab,kw OR 'maldevelopment':ti,ab,kw OR 'deprivation':ti,ab,kw OR 'offence':ti,ab,kw OR 'neglect':ti,ab,kw OR 'IPV':ti,ab,kw OR 'stigma':ti,ab,kw OR 'psychological discrimination':ti,ab,kw OR 'social discrimination':ti,ab,kw OR 'perceived discrimination':ti,ab,kw OR 'prejudice':ti,ab,kw OR 'stigmatization':ti,ab,kw OR 'stigmatisation':ti,ab,kw OR 'stigmatized':ti,ab,kw OR 'stigmatised':ti,ab,kw OR 'discriminate':ti,ab,kw OR 'discrimination':ti,ab,kw OR 'discriminating':ti,ab,kw OR 'discriminated':ti,ab,kw OR 'oppress':ti,ab,kw OR 'oppressed':ti,ab,kw OR 'oppressing':ti,ab,kw OR 'oppression':ti,ab,kw OR 'transphobia':ti,ab,kw OR 'transphobic':ti,ab,kw OR 'prejudice':ti,ab,kw OR 'stereotype':ti,ab,kw OR 'stereotyping':ti,ab,kw OR 'stereotyped attitudes':ti,ab,kw OR 'shame':ti,ab,kw OR 'disrespect':ti,ab,kw OR 'coercive control':ti,ab,kw OR 'forced procedures':ti,ab,kw OR 'interpersonal violence':ti,ab,kw OR 'psychological violence':ti,ab,kw

**CINAHL**

**Concept 1: TGD population**

(MM "Sexual and Gender Minorities+") OR "sexual and gender minorities" OR (MM "Conversion Therapy") OR (MM "Gender-Nonconforming Persons+") OR (MM "Cross-Dressing") OR (MM "Cross-dressers") OR (MM "Transsexuals") OR ((AB "achout") OR (AB "AFAB") OR (AB "agender") OR (AB "akavaine") OR (AB "AMAB") OR (AB "aravanis") OR (AB "assigned-female-at-birth") OR (AB "assigned-female-sex") OR (AB "assigned-male-at-birth") OR (AB "assigned-male-sex") OR (AB "bakla") OR (AB "bantut") OR (AB "berdache") OR (AB "bigender") OR (AB "bi-gender") OR (AB "biological-female") OR (AB "biological-male") OR (AB "biological-man") OR (AB "biological-men") OR (AB "biological-woman") OR (AB "biological-women") OR (AB "brotherboys") OR (AB "brother-boys") OR (AB "cross-dress") OR (AB "dissident-gender") OR (AB "faafafine") OR (AB "fakafifine") OR (AB "female-to-male") OR (AB "gender-affirm") OR (AB "gender-atypical") OR (AB "atypical-gender") OR (AB "gender-bend") OR (AB "gender-binary") OR (AB "gender-change") OR (AB "gender-disorder") OR (AB "gender-diverse") OR (AB "genderdiverse") OR (AB "gender-dysphoria") OR (AB "gender-expansive") OR (AB "genderexpansive") OR (AB "genderfluid") OR (AB "gender-fluid") OR (AB "gender-identity-disorder") OR (AB "gender-inclusive") OR (AB "gender-incongruent") OR (AB "gender-minority") OR (AB "gender-neutral") OR (AB "gender-orientation") OR (AB "genderqueer") OR (AB "genderquestioning") OR (AB "gender-questioning") OR (AB "gender-reassign") OR (AB "gender-re-assign") OR (AB "gender-variant") OR (AB "hijra") OR (AB "irahuhua") OR (AB "intersex") OR (AB "irawhiti") OR (AB "kathoey") OR (AB "kathoy") OR (AB "katoey") OR (AB "khanith") OR (AB "khwaja sira") OR (AB "kothi") OR (AB "koti") OR (AB "leiti") OR (AB "machi") OR (AB "mah") OR (AB "mahuvahine") OR (AB "male-to-female") OR (AB "mak-nyah") OR (AB "maknyah") OR (AB "metis") OR (AB "minority-gender") OR (AB "muxe") OR (AB "nadleehi") OR (AB "non-binary") OR (AB "nonbinary") OR (AB "non-conforming") OR (AB "nonconforming") OR (AB "paknyah") OR (AB "palopa") OR (AB "phuying-kham-phet") OR (AB "pinapinaaine") OR (AB "queer") OR (AB "sao-praphet-song") OR (AB "selrata") OR (AB "sex-affirming") OR (AB "sex-change") OR (AB "sex-reassignment") OR (AB "sex-re-assignment") OR (AB "sexual-dissident") OR (AB "sexual-dysphoria") OR (AB "sistergirls") OR (AB "sister-girls") OR (AB "sworn-virgin") OR (AB "tahine") OR (AB "takataapui") OR (AB "tangata-ira-tane") OR (AB "tangata-ira-wahine") OR (AB "third-gender") OR (AB "third-spirit") OR (AB "transbodies") OR (AB "trans-bodies") OR (AB "transboy") OR (AB "trans-boy") OR (AB "trans-experience") OR (AB "transfeminine") OR (AB "trans-feminine") OR (AB "transgender") OR (AB "trans-gender") OR (AB "transgirl") OR (AB "trans-girl") OR (AB "trans-identity") OR (AB "transidentity") OR (AB "transmale") OR (AB "trans-male") OR (AB "transman") OR (AB "trans-man") OR (AB "transmen") OR (AB "trans-men") OR (AB "transmasculine") OR (AB "trans-masculine") OR (AB "trans-people") OR (AB "transpeople") OR (AB "trans-person") OR (AB "trans-persons") OR (AB "transperson") OR (AB "transpersons") OR (AB "transpinay") OR (AB "transpinoy") OR (AB "trans-selves") OR (AB "transsexual") OR (AB "trans-sexual") OR (AB "transvestite") OR (AB "transvisibility") OR (AB "trans-visibility") OR (AB "transwoman") OR (AB "trans-woman") OR (AB "travesti") OR (AB "two-spirit") OR (AB "2-spirit") OR (AB "vakasalewalewa") OR (AB "waria") OR (AB "whakawahine") OR (AB "xanith") NOT ((AB "male-to-female ratio") OR (AB "male-to-female ratios") OR (AB "female-to-male ratio") OR (AB "female-to-male ratios") OR (AB "transfemoral")))

**Concept 2: stigma/discrimination/violence**

(MM "Violence+") OR (MM "Intimate Partner Violence") OR (MM "Gender-Based Violence") OR (MM "Dating Violence") OR (MM "Exposure to Violence") OR (MM "Stigma") OR (MM "Discrimination+") OR (MM "Perceived Discrimination") OR (MM "Transphobia") OR (MM "Prejudice+") OR (MH "Stereotyping") OR (MH "Implicit Bias") OR (AB "violence") OR (AB "gender-based violence") OR (AB "abuse") OR (AB "rape") OR (AB "forced sex") OR (AB "coerced sex") OR (AB "molestation") OR (AB "psychological harm") OR (AB "maldevelopment") OR (AB "deprivation") OR (AB "offence") OR (AB "neglect") OR (AB "IPV") OR (AB "stigma") OR (AB "psychological discrimination") OR (AB "social discrimination") OR (AB "perceived discrimination") OR (AB "prejudice") OR (AB "stigmatization") OR (AB "stigmatisation") OR (AB "stigmatized") OR (AB "stigmatised") OR (AB "discriminate") OR (AB "discrimination") OR (AB "discriminating") OR (AB "discriminated") OR (AB "oppress") OR (AB "oppressed") OR (AB "oppressing") OR (AB "oppression") OR (AB "transphobia") OR (AB "transphobic") OR (AB "prejudice") OR (AB "stereotype") OR (AB "stereotyping") OR (AB "stereotyped attitudes") OR (AB "shame") OR (AB "disrespect") OR (AB "coercive control") OR (AB "forced procedures") OR (AB "interpersonal violence") OR (AB "psychological violence")

**PsycInfo**

**Concept 1: TGD population**

(MM "Sexual and Gender Minorities+") OR "sexual and gender minorities" OR (MM "Conversion Therapy") OR (MM "Gender-Nonconforming Persons+") OR (MM "Cross-Dressing") OR (MM "Cross-dressers") OR (MM "Transsexuals") OR ((AB "achout") OR (AB "AFAB") OR (AB "agender") OR (AB "akavaine") OR (AB "AMAB") OR (AB "aravanis") OR (AB "assigned-female-at-birth") OR (AB "assigned-female-sex") OR (AB "assigned-male-at-birth") OR (AB "assigned-male-sex") OR (AB "bakla") OR (AB "bantut") OR (AB "berdache") OR (AB "bigender") OR (AB "bi-gender") OR (AB "biological-female") OR (AB "biological-male") OR (AB "biological-man") OR (AB "biological-men") OR (AB "biological-woman") OR (AB "biological-women") OR (AB "brotherboys") OR (AB "brother-boys") OR (AB "cross-dress") OR (AB "dissident-gender") OR (AB "faafafine") OR (AB "fakafifine") OR (AB "female-to-male") OR (AB "gender-affirm") OR (AB "gender-atypical") OR (AB "atypical-gender") OR (AB "gender-bend") OR (AB "gender-binary") OR (AB "gender-change") OR (AB "gender-disorder") OR (AB "gender-diverse") OR (AB "genderdiverse") OR (AB "gender-dysphoria") OR (AB "gender-expansive") OR (AB "genderexpansive") OR (AB "genderfluid") OR (AB "gender-fluid") OR (AB "gender-identity-disorder") OR (AB "gender-inclusive") OR (AB "gender-incongruent") OR (AB "gender-minority") OR (AB "gender-neutral") OR (AB "gender-orientation") OR (AB "genderqueer") OR (AB "genderquestioning") OR (AB "gender-questioning") OR (AB "gender-reassign") OR (AB "gender-re-assign") OR (AB "gender-variant") OR (AB "hijra") OR (AB "irahuhua") OR (AB "intersex") OR (AB "irawhiti") OR (AB "kathoey") OR (AB "kathoy") OR (AB "katoey") OR (AB "khanith") OR (AB "khwaja sira") OR (AB "kothi") OR (AB "koti") OR (AB "leiti") OR (AB "machi") OR (AB "mah") OR (AB "mahuvahine") OR (AB "male-to-female") OR (AB "mak-nyah") OR (AB "maknyah") OR (AB "metis") OR (AB "minority-gender") OR (AB "muxe") OR (AB "nadleehi") OR (AB "non-binary") OR (AB "nonbinary") OR (AB "non-conforming") OR (AB "nonconforming") OR (AB "paknyah") OR (AB "palopa") OR (AB "phuying-kham-phet") OR (AB "pinapinaaine") OR (AB "queer") OR (AB "sao-praphet-song") OR (AB "selrata") OR (AB "sex-affirming") OR (AB "sex-change") OR (AB "sex-reassignment") OR (AB "sex-re-assignment") OR (AB "sexual-dissident") OR (AB "sexual-dysphoria") OR (AB "sistergirls") OR (AB "sister-girls") OR (AB "sworn-virgin") OR (AB "tahine") OR (AB "takataapui") OR (AB "tangata-ira-tane") OR (AB "tangata-ira-wahine") OR (AB "third-gender") OR (AB "third-spirit") OR (AB "transbodies") OR (AB "trans-bodies") OR (AB "transboy") OR (AB "trans-boy") OR (AB "trans-experience") OR (AB "transfeminine") OR (AB "trans-feminine") OR (AB "transgender") OR (AB "trans-gender") OR (AB "transgirl") OR (AB "trans-girl") OR (AB "trans-identity") OR (AB "transidentity") OR (AB "transmale") OR (AB "trans-male") OR (AB "transman") OR (AB "trans-man") OR (AB "transmen") OR (AB "trans-men") OR (AB "transmasculine") OR (AB "trans-masculine") OR (AB "trans-people") OR (AB "transpeople") OR (AB "trans-person") OR (AB "trans-persons") OR (AB "transperson") OR (AB "transpersons") OR (AB "transpinay") OR (AB "transpinoy") OR (AB "trans-selves") OR (AB "transsexual") OR (AB "trans-sexual") OR (AB "transvestite") OR (AB "transvisibility") OR (AB "trans-visibility") OR (AB "transwoman") OR (AB "trans-woman") OR (AB "travesti") OR (AB "two-spirit") OR (AB "2-spirit") OR (AB "vakasalewalewa") OR (AB "waria") OR (AB "whakawahine") OR (AB "xanith") NOT ((AB "male-to-female ratio") OR (AB "male-to-female ratios") OR (AB "female-to-male ratio") OR (AB "female-to-male ratios") OR (AB "transfemoral")))

**Concept 2: stigma/discrimination/violence**

(MM "Violence+") OR (MM "Intimate Partner Violence") OR (MM "Gender-Based Violence") OR (MM "Dating Violence") OR (MM "Exposure to Violence") OR (MM "Stigma") OR (MM "Discrimination+") OR (MM "Perceived Discrimination") OR (MM "Transphobia") OR (MM "Prejudice+") OR (MH "Stereotyping") OR (MH "Implicit Bias") OR (AB "violence") OR (AB "gender-based violence") OR (AB "abuse") OR (AB "rape") OR (AB "forced sex") OR (AB "coerced sex") OR (AB "molestation") OR (AB "psychological harm") OR (AB "maldevelopment") OR (AB "deprivation") OR (AB "offence") OR (AB "neglect") OR (AB "IPV") OR (AB "stigma") OR (AB "psychological discrimination") OR (AB "social discrimination") OR (AB "perceived discrimination") OR (AB "prejudice") OR (AB "stigmatization") OR (AB "stigmatisation") OR (AB "stigmatized") OR (AB "stigmatised") OR (AB "discriminate") OR (AB "discrimination") OR (AB "discriminating") OR (AB "discriminated") OR (AB "oppress") OR (AB "oppressed") OR (AB "oppressing") OR (AB "oppression") OR (AB "transphobia") OR (AB "transphobic") OR (AB "prejudice") OR (AB "stereotype") OR (AB "stereotyping") OR (AB "stereotyped attitudes") OR (AB "shame") OR (AB "disrespect") OR (AB "coercive control") OR (AB "forced procedures") OR (AB "interpersonal violence") OR (AB "psychological violence")

**Cochrane Central**

MeSH descriptor: [Transgender Persons] explode all trees

**LILACS**

**Concept 1: TGD population**

(mh:m01.270.988* OR mh:("Transsexualism" OR "Gender Dysphoria" OR "Health Services for Transgender Persons" OR "Sex Reassignment Procedures" OR "Sex Reassignment Surgery" OR "Transvestism") OR (achout OR afab OR agender OR "akava'ine" OR amab OR aravanis OR "assigned female" OR "assigned male" OR bakla OR bantut OR berdache OR bigender* OR bi-gender* OR "biological female" OR "biological male" OR "biological females" OR "biological males" OR "biological man" OR "biological men" OR "biological woman" OR "biological women" OR brotherboys OR brother-boys OR "cross dressing" OR "cross dressed" OR "dissident gender" OR "dissident genders" OR "fa'afafine" OR fakafifine OR "female to male" OR "female to males" OR "gender affirming" OR "gender affirmation" OR "gender atypical" OR "gender atypicality" OR "atypical gender" OR "atypicality gender" OR "gender bending" OR "gender bendingand" OR "gender binaries" OR "gender binary" OR "gender change" OR "gender changes" OR "gender disorder" OR "gender disorders" OR "gender diverse" OR "gender diversity" OR genderdivers* OR "gender dysphoric" OR "gender dysphoria" OR "gender expansive" OR genderexpans* OR genderfluid OR "gender fluid" OR "gender identity" OR "gender inclusive" OR "gender incongruent" OR "gender incongruence" OR "gender minority" OR "gender minorities" OR "gender neutral" OR "gender orientation" OR "gender oriented" OR "gender orientations" OR genderqueer* OR genderquestioning OR "gender questioning" OR "gender reassigning" OR "gender reassignment" OR "gender reassigned" OR "gender re-assignment" OR "gender re-assigned" OR "gender re-assigning" OR "gender variant" OR "gender variants" OR hijra* OR irahuhua OR intersex* OR irawhiti OR kathoey OR kathoy OR katoey OR khanith OR khwaja-sira OR khwaja-sara OR kothi OR koti OR leiti OR machi OR mah OR mahuvahine OR "male to female" OR "male to females" OR mak-nyah OR maknyah OR metis OR minority-gender OR muxe OR nadleehi OR non-binar* OR nonbinar* OR non-conform* OR nonconform* OR paknyah OR palopa OR phuying-kham-phet OR pinapinaaine OR queer* OR sao-praphet-song OR selrata OR "sex affirming" OR "sex affirmation" OR "sex change" OR "sex changes" OR "sex reassigning" OR "sex reassignment" OR "sex reassignment" OR "sex re-assignment" OR "sex re-assigned" OR "sex re-assigning" OR "sexual dissident" OR "sexual dysphoric" OR "sexual dysphoria" OR sistergirls OR sister-girls OR "Sworn virgins" OR tahine OR takataapui OR tangata-ira-tane OR tangata-ira-wahine OR "third gender" OR "third spirit" OR transbod* OR "trans bodies" OR transboy* OR "trans boy" OR "trans boys" OR "trans experience" OR "trans experiences" OR transfem* OR "trans female" OR transgender* OR "trans gender" OR "trans genders" OR transgirl* OR "trans girl" OR "trans girls" OR "trans identity" OR "trans identities" OR "trans identifying" OR "trans identified" OR "trans identical" OR "trans identification" OR transident* OR transmale* OR "trans male" OR transman OR "trans man" OR transmen OR "trans men" OR transmasculin* OR "trans masculine" OR "trans people" OR transpeople OR "trans person" OR "trans persons" OR transperson OR transpersons OR transpinay* OR transpinoy* OR trans-selv* OR transsex* OR "trans sexual" OR "trans sexualism" OR "trans sexuality" OR "trans sexualities" OR "trans sexuals" OR "trans sex" OR transvest* OR transvisib* OR trans-visib* OR transwom* OR "trans woman" OR "trans women" OR travest* OR "two spirit" OR 2-spirit* OR lgbt* OR vakasalewalewa OR waria OR whakawahine OR xanith)) AND NOT ((("male to female" OR "males to female" OR "male to females" OR "males to females" OR "female to male" OR "females to male" OR "female to males" OR "females to males") AND ratio*) OR transfemoral)

**Concept 2: stigma/discrimination/violence**

ti,ab,mh:("social stigmas" OR "Perceived Discrimination" OR "Social Discrimination" OR "Violence" OR "Gender-based Violence" OR "Stereotyping")

**Organizational websites searched as part of the grey literature search strategy:**

Asia Pacific Transgender Network. Asia Pacific. https://weareaptn.org/
Callen-Lorde. New York, NY, USA. https://callen-lorde.org/
Center of Excellence for Transgender Health. San Francisco, CA, USA. https://prevention.ucsf.edu/transhealth
Chase Brexton. Baltimore, MD, USA. https://chasebrexton.org/
Chulalongkorn University (Center of Excellence in Transgender Health). Thailand. https://www.chula.ac.th/en/, https://www.chula.ac.th/en/highlight/47996/
Fenway Health. Boston, MA, USA. https://fenwayhealth.org/
Gender Wellbeing Clinic. Malta. https://genderwellbeingclinic.business.site/
GenderGP: Online Transgender Clinic. Worldwide, online. https://www.gendergp.com/
genderHealthCare. Utrench, Netherlands. https://genderhealthcare.com/
Ghent University Hospital (Center of Sexology and Gender). Belgium. https://www.crcg.ugent.be/, https://epath.eu/wp-content/uploads/2014/07/Transgender-health-care-in-Belgium_Els-Elaut_20150313.pdf
Ghent University Hospital (Department of Endocrinology). Belgium. https://www.ugent.be/ge/izp/en/research/research-themes/endocrinology.htm
Howard Brown Health. Chicago, IL. https://howardbrown.org/
ILGA. Brussels, Belgium. https://www.ilga-europe.org/about-us/
Indigo Gender Service. Manchester, UK. https://indigogenderservice.uk/about-us

Institute of HIV Testing and Innovation (IHRI). Thailand. https://ihri.org/
International Lesbian, Gay, Bisexual, Trans and Intersex Association (IGLA Europe). Brussels, Belgium. https://www.ilga-europe.org/
Legacy Community Health. Houston, TX, USA. https://www.legacycommunityhealth.org/
Lyon Martin Community Health. San Francisco, CA, USA. https://lyon-martin.org/
Mazzoni Center. Philadelphia, PA, USA. https://www.mazzonicenter.org/
Mitr Clinic (Fenway Health). Hyderabad, India. https://www.safezindagi.in/mitr-clinic
National Center for Transgender Equality. Washington DC, USA. https://transequality.org/
Okayama University (Hospital Gender Center). Okayama, Japan. https://www.okayama-u.ac.jp/user/hospital/en/index186.html
Soweto Transgender Care. Soweto, Johannesburg, South Africa. https://www.sowetotransgender.co.za/#:~:text=For%20now%2C%20Soweto%20Transgender%20Care,surgeries%2C%20and%20other%20supportive%20services.
The Swedish Federation for Lesbian, Gay, Bisexual, Transgender, Queer and Intersex Rights (RSFL). Sweden. https://www.rfsl.se/en/organisation/vard-for-transpersoner/transvaard/
Trans United Netherland (trans-led clinic). Amsterdam, Netherlands. https://transunitedeurope.eu/
TransCare. Athens, Greece. https://transcare-project.eu/
Transgender EU (TGEU). Berlin, Germany. https://tgeu.org/about-us/
University of Witwaterand (WITS RHI). Johannesburg, South Africa. https://www.wrhi.ac.za/media/detail/first-dedicated-clinics-to-open-for-transgender-care-under-key-populat
Whitman-Walker Health. Washington, DC, USA. https://www.whitman-walker.org/
Williams Institute, UCLA School of Law. Los Angeles, CA, USA. https://williamsinstitute.law.ucla.edu/, https://williamsinstitute.law.ucla.edu/subpopulations/transgender-people/, https://williamsinstitute.law.ucla.edu/visualization/lgbt-stats/?topic=LGBT#density

**Appendix B. Included articles**

| First Author | Publication Year | Article Title | Sample Size | Country | Study Design | Anticipated Stigma | Internalized Stigma | Perceived Stigma | Experienced Stigma | Experienced Discrimination | Resilience |
|---|---|---|---|---|---|---|---|---|---|---|---|
| Anderson | 2021 | Characteristics of sexual and gender minority caregivers of people with dementia | 38 | U.S. | Survey | | | X | | | |
| Azhar | 2022 | Associations between HIV stigma, gender, and depression among people living with HIV in Hyderabad, India | 50 | India | Survey | | | | X | | |
| Barr | 2022 | Posttraumatic stress in the trans community: The roles of anti-transgender bias, non-affirmation, and internalized transphobia | 575 | U.S. | Survey | | X | | | | |
| Basar | 2016 | Perceived Discrimination, Social Support, and Quality of Life in Gender Dysphoria | 116 | Turkey | Survey | | | X | | | |
| Basar | 2016 | Resilience in Individuals with Gender Dysphoria: Association with Perceived Social Support and Discrimination | 116 | Turkey | Survey | | | | | | X |
| Bauermeister | 2016 | Psychosocial Disparities Among Racial/Ethnic Minority Transgender Young Adults and Young Men Who Have Sex with Men Living in Detroit | 26 | U.S. | Survey | | | | | X | |
| Boza | 2014 | Gender-Related Victimization, Perceived Social Support, and Predictors of Depression Among Transgender Australians | 243 | Australi a | Survey | | | | | X | |
| Bretherton | 2021 | The Health and Well-Being of Transgender Australians: A National Community Survey | 927 | Australi a | Survey | | | | | X | |
| Budhwani | 2017 | Transgender female sex workers' HIV knowledge, experienced stigma, and condom use in 87the Dominican Republic | 78 | Dominic an Republi c | Survey | | | | | X | |

| | | | | | | | | | | |
|---|---|---|---|---|---|---|---|---|---|---|
| Busby | 2020 | Suicide risk among gender and sexual minority college students: The roles of victimization, discrimination, connectedness, and identity affirmation | 87 | U.S. | Survey | | | X | | |
| Caceres | 2022 | Examining the Associations of Gender Minority Stressors with Sleep Health in Gender Minority Individuals | 279 | U.S. | Longitudinal | | | X | | |
| Casey | 2019 | Discrimination in the United States: Experiences of lesbian, gay, bisexual, transgender, and queer Americans | 86 | U.S. | Survey | | | | | X |
| De Mattos Russo Rafael | 2021 | Prevalence and factors associated with suicidal behavior among trans women in Rio de Janeiro, Brazil | 345 | Brazil | Survey | | | | | X |
| Fredriksen-Goldsen | 2013 | Physical and Mental Health of Transgender Older Adults: An At-Risk and Underserved Population | 174 | U.S. | Survey | | X | X | | |
| Fritz | 2016 | Social Demography of Health Seeking Experiences Among Transgender African Americans | 253 | U.S. | Survey/Secondary Data Analysis | | | | | X |
| Gamarel | 2014 | Gender minority stress, mental health, and relationship quality: A dyadic investigation of transgender women and their cisgender male partners | 191 | U.S. | Survey | | X | | X | |
| Gamarel | 2016 | Minority Stress, Smoking Patterns, and Cessation Attempts: Findings From a Community-Sample of Transgender Women in the San Francisco Bay Area | 241 | U.S. | Survey | | | | | X |
| Glick | 2018 | The Role of Discrimination in Care Postponement Among Trans-Feminine Individuals in the U.S. National Transgender Discrimination Survey | 2248 | U.S. | Survey/Secondary Data Analysis | | | | | X |
| House | 2011 | Interpersonal trauma and discriminatory events as predictors of suicidal and nonsuicidal self-injury in gay, lesbian, bisexual, and transgender persons | 164 | U.S. | Survey | | | | | X |
| Hsiang | 2022 | Prevalence and Correlates of Substance Use and Associations with HIV-Related Outcomes | 629 | U.S. | Survey/Secondary | | | | | X |

| | | | | | | | | | | |
|---|---|---|---|---|---|---|---|---|---|---|
| | | Among Trans Women in the San Francisco Bay Area | | | Data Analysis | | | | | |
| Hughto | 2021 | Opioid pain medication misuse, concomitant substance misuse, and the unmet behavioral health treatment needs of transgender and gender diverse adults | 562 | U.S. | Survey | | | | | X |
| Hughto | 2022 | Victimization Within and Beyond the Prison Walls: Latent Profile Analysis of Transgender and Gender Diverse Adults | 574 | U.S. | Survey | | | | X | X |
| Jaggi | 2018 | Gender Minority Stress and Depressive Symptoms in Transitioned Swiss Transpersons | 143 | Switzerl and | Survey | X | | | | X |
| Kachen | 2021 | Health Care Access and Utilization by Transgender Populations: A United States Transgender Survey Study | 2771 5 | U.S. | Survey/Se condary Data Analysis | | | | | X |
| Kachen | 2022 | Creating a minority stress index to examine mental health impacts of discrimination among transgender and gender nonbinary adults | 2771 5 | U.S. | Survey/Se condary Data Analysis | | | | | X |
| Kaplan | 2016 | HIV prevalence and demographic determinants of condomless receptive anal intercourse among trans feminine individuals in Beirut, Lebanon | 53 | Lebanon | Survey | | | | | X |
| Kattari | 2015 | Racial and Ethnic Differences in Experiences of Discrimination in Accessing Social Services Among Transgender/Gender-Nonconforming People | 6451 | U.S. | Survey/Se condary Data Analysis | | | | | X |
| Kattari | 2015 | Differences Across Age Groups in Transgender and Gender Non- Conforming People's Experiences of Health Care Discrimination, Harassment, and Victimization | 5006 | U.S. | Survey/Se condary Data Analysis | | | | | X |
| Kattari | 2016 | Policing Gender Through Housing and Employment Discrimination: Comparison of Discrimination Experiences of Transgender and Cisgender LGBQ Individuals | 148 | U.S. | Survey | | | | | X |
| Kattari | 2017 | Differences in Experiences of Discrimination in Accessing Social Services Among | 6456 | U.S. | Survey/Se condary | | | | | X |

| | | | | | | | | | | |
|---|---|---|---|---|---|---|---|---|---|---|
| | | Transgender/ Gender Nonconforming Individuals by (Dis)Ability | | | Data Analysis | | | | | |
| Kcomt | 2020 | Association of transphobic discrimination and alcohol misuse among transgender adults: results from the US transgender survey | 27715 | U.S. | Survey/Secondary Data Analysis | | | | | X |
| Kcomt | 2020 | Use of cigarettes and e-cigarettes/vaping among transgender people: results from the 2015 US transgender survey | 27715 | U.S. | Survey/Secondary Data Analysis | | | | | X |
| Kidd | 2019 | Understanding predictors of improvement in risky drinking in a U.S. multi-site, longitudinal cohort study of transgender individuals: Implications for culturally-tailored prevention and treatment efforts | 286 | U.S. | Longitudinal | | X | | | |
| Klemmer | 2021 | Transphobia-Based Violence, Depression, and Anxiety in Transgender Women: The Role of Body Satisfaction | 233 | U.S. | Survey | | | | | X |
| Lacombe-Duncan | 2019 | The HIV Care Cascade Among Transgender Women with HIV in Canada: A Mixed-Methods Study | 50 | Canada | Survey/Secondary Data Analysis | | | | | X |
| Lacombe-Duncan | 2019 | Gender-affirming healthcare experiences and medical transition among transgender women living with HIV: a mixed-methods study | 48 | Canada | Survey/Secondary Data Analysis | | | | | X |
| Lee | 2021 | Transgender Adults' Public Bathroom-Related Stressors and Their Association with Depressive Symptoms: A Nationwide Cross-Sectional Study in South Korea | 557 | South Korea | Survey | X | | | | |
| Lee | 2022 | Does Discrimination Affect Whether Transgender People Avoid or Delay Healthcare?: A Nationwide Cross-Sectional Survey in South Korea | 244 | South Korea | Survey | | | | | X |
| Leite | 2021 | Associations between gender-based discrimination and medical visits and HIV | 864 | Brazil | Survey | | | | | X |

| | | | | | | | | | | | |
|---|---|---|---|---|---|---|---|---|---|---|---|
| | | testing in a large sample of transgender women in northeast Brazil | | | | | | | | | |
| Lerner | 2020 | More than an Apple a Day: Factors Associated with Avoidance of Doctor Visits Among Transgender, Gender Nonconforming, and Nonbinary People in the USA | 21930 | U.S. | Survey/Secondary Data Analysis | | | | | X | |
| Lerner | 2021 | Having to 'Hold It': Factors That influence the Avoidance of Using Public Bathrooms Among Transgender People | 25694 | U.S. | Survey/Secondary Data Analysis | X | | | | | |
| Levine | 2022 | Associations Between Healthcare Experiences, Mental Health Outcomes, and Substance Use Among Transgender Adults | 18890 | U.S. | Survey/Secondary Data Analysis | | | | | X | |
| Lewis | 2019 | Transgender/gender nonconforming adults' worries and coping actions related to discrimination: Relevance to pharmacist care | 316 | U.S. | Survey | X | | | | X | |
| Logie | 2019 | Syndemic Experiences, Protective Factors, and HIV | 137 | Jamaica | Survey | | | | | | X |
| Lozano-Verduzco | 2021 | Transgender individuals in Mexico: exploring characteristics and experiences of discrimination and violence | 148 | Mexico | Survey | | | | | X | X |
| Luz | 2022 | Associations of Discrimination, Violence, and Resilience with Depressive Symptoms Among Transgender Women in Rio de Janeiro, Brazil: A Cross-Sectional Analysis | 489 | Brazil | Survey/Secondary Data Analysis | | | X | | | |
| Marshall | 2016 | Prevalence and Correlates of Lifetime Suicide Attempts Among Transgender Persons in Argentina | 482 | Argentina | Survey/Secondary Data Analysis | | X | | | | |
| McDowell | 2019 | Risk and protective factors for mental health morbidity in a community sample of female-to-male trans-masculine adults | 150 | United States | Survey | | | X | | | |
| Messinger | 2022 | Intimate Partner Violence Help-Seeking in the U.S. Transgender Survey | 15198 | U.S. | Survey/Secondary Data Analysis | | | | | X | |

| | | | | | | | | | | |
|---|---|---|---|---|---|---|---|---|---|---|
| MezaLazaro | 2021 | Determinants of Mental Health Outcomes Among Transgender Latinas: Minority Stress and Resilience Processes | 424 | U.S. | Survey/Secondary Data Analysis | | | | | X |
| Milner | 2019 | Sex work, social support, and stigma: Experiences of transgender women in the Dominican Republic | 291 | Dominican Republic | Survey | | | | X | X |
| Nematollahi | 2022 | Discrimination, Violence, and Suicide in Transgender Women in Iran | 127 | Iran | Survey | | | | X | |
| Nemoto | 2011 | Social Support, Exposure to Violence and Transphobia, and Correlates of Depression Among Male-to-Female Transgender Women With a History of Sex Work | 573 | U.S. | Survey | | | | X | X |
| Palve | 2018 | Health Issues Among Transgenders in Urban pondicherry | 121 | India | Survey | | | | | |
| Parr | 2019 | Heterogeneity of Transgender Identity Nonaffirmation Microaggressions and Their Association with Depression Symptoms and Suicidality Among Transgender Persons | 182 | U.S. | Survey/ Focus Group | | | | | X |
| Poteat | 2017 | HIV prevalence and behavioral and psychosocial factors among transgender women and cisgender men who have sex with men in 8 African countries: A cross-sectional analysis | 937 | 8 African Sites | Survey | | | | X | |
| Rafael | 2021 | Prevalence and factors associated with suicidal behavior among trans women in Rio de Janeiro, Brazil | 345 | Brazil | Survey | | | | | X |
| Ralston | 2022 | Mental health and marginalization stress in transgender and gender diverse adults: Differences between urban and non-urban experiences | 225 | U.S. | Survey | | X | | | X |
| Rodriguez-Madera | 2017 | Experiences of Violence Among Transgender Women in Puerto Rico: An Underestimated Problem | 59 | U.S. | Survey | | | | | X |
| Romanelli | 2018 | Examining Mechanisms and Moderators of the Relationship Between Discriminatory Health | 4190 | U.S. | Survey/Secondary | | | | | X |

| | | | | | | | | | | |
|---|---|---|---|---|---|---|---|---|---|---|
| | | Care Encounters and Attempted Suicide Among U.S. Transgender Help-Seekers | | | Data Analysis | | | | | |
| Romanelli | 2020 | Patterns of Healthcare Discrimination Among Transgender Help-Seekers | 2354 1 | U.S. | Survey/Se condary Data Analysis | | | | | X |
| Rood | 2015 | Predictors of Suicidal Ideation in a Statewide Sample of Transgender Individuals | 350 | U.S. | Survey | | | | | X |
| Rosentel | 2021 | Black Transgender Women and the School-to-Prison Pipeline: Exploring the Relationship Between Anti-trans Experiences in School and Adverse Criminal-Legal System Outcomes | 138 | U.S. | Survey | X | | | | X |
| Rotondi | 2011 | Depression in Male-to-Female Transgender Ontarians: Results from the Trans PULSE Project | 191 | Canada | Survey | | | | | X |
| Rouhani | 2021 | Resilience among Cisgender and Transgender Women in Street-Based Sex Work in Baltimore, Maryland | 42 | U.S. | Longitudi nal | | | | | X |
| Ruggs | 2015 | Workplace 'Trans' Actions - How Organizations, Coworkers, and Individual Openness Influence Perceived Gender Identity Discrimination | 118 | U.S. | Survey | | X | | | |
| Salas-Espinoza | 2017 | HIV Prevalence and Risk Behaviors in Male to Female (MTF) Transgender Persons in Tijuana, Mexico | 100 | Mexico | Survey | X | X | | | |
| Salazar | 2017 | Contextual, Experiential, and Behavioral Risk Factors Associated with HIV Status: a Descriptive Analysis of Transgender Women Residing in Atlanta, Georgia | 92 | U.S. | Survey | | X | | | |
| Scandurra | 2020 | The Italian validation of the gender minority stress and resilience measure | 203 | Italy | Survey | | X | | | |
| Scheim | 2016 | Inequities in access to HIV prevention services for transgender men: results of a global survey of men who have sex with men | 69 | Global | Survey | | | | | X |
| Scheim | 2017 | HIV-Related Sexual Risk Among Transgender Men Who Are Gay, Bisexual, or Have Sex With Men | 158 | Canada | Survey | | | | | X |

| | | | | | | | | | | |
|---|---|---|---|---|---|---|---|---|---|---|
| Schweizer | 2022 | Discrimination and Risky Sexual Behavior, Substance Use, and Suicidality among Transgender Individuals | 350 | U.S. | Survey/Secondary Data Analysis | | | | | X |
| Seelman | 2016 | Transgender Adults' Access to College Bathrooms and Housing and the Relationship to Suicidality | 2325 | U.S. | Survey/Secondary Data Analysis | | | | | X |
| Seelman | 2020 | Trans men’s access to knowledgeable providers and their experiences in health care settings: Differences by demographics, mental health, and degree of being 'out' to providers | 7950 | U.S. | Survey/Secondary Data Analysis | | | | | X |
| Sevelius | 2014 | Correlates of antiretroviral adherence and viral load among transgender women living with HIV | 59 | U.S. | Survey | | | | | X |
| Shah | 2018 | Challenges faced by marginalized communities such as transgender in Pakistan | 189 | Pakistan | Survey | | | | | X |
| She | 2021 | Mental health service utilisation among transgender women sex workers who are at risk of mental health problems in Shenyang, China: An application of minority stress theory | 199 | China | Survey | X | | | | |
| She | 2021 | Impact of minority stress and poor mental health on sexual risk behaviors among transgender women sex workers in Shenyang, China | 204 | China | Survey | | | | X | |
| Shires | 2015 | Factors Associated with Health Care Discrimination Experiences among a national Sample Female-to -Male Transgender Individuals | 1711 | U.S. | Survey/Secondary Data Analysis | | | | | X |
| Siamisang | 2022 | High-risk behaviors and factors for HIV and sexually transmitted infections among transgender people in Gaborone, Botswana: results from a national survey | 12 | Botswana | Survey/Secondary Data Analysis | | | | | X |
| Silva | 2021 | Factors associated with suicidal ideation among transvestites and transsexuals assisted by non-governmental organizations | 58 | Brazil | Cross-Sectional Study | | | | X | |
| Silva | 2022 | Transgender parenthood, participation in children's lives, and association with | 670 | Brazil | Secondary Analysis | | | | X | X |

| | | | | | | | | | | | |
|---|---|---|---|---|---|---|---|---|---|---|---|
| | | discrimination experiences: An exploratory study | | | | | | | | | |
| Sutherland | 2021 | Exploring factors contributing to care-seekers' level of discomfort discussing a transgender identity in a health care setting | 344 | U.S. | Survey/Secondary Data Analysis | | | | | X | |
| Tabaac | 2018 | Discrimination, mental health, and body image among transgender and gender-non-binary individuals: Constructing a multiple mediational path model | 78 | U.S. | Survey | | | | | X | |
| Thompson-Blum | 2021 | Experiences of Transgender Participants in Emergency Departments: Findings from the OutLook Study | 112 | Canada | Survey | | | X | | | |
| Valente | 2020 | Stigmatization, Resilience, and Mental Health Among a Diverse Community Sample of Transgender and Gender Nonbinary Individuals in the U.S | 330 | U.S. | Longitudinal | | | | | X | X |
| Valente | 2022 | Prospective relationships between stigma, mental health, and resilience in a multi-city cohort of transgender and nonbinary individuals in the United States, 2016-2019 | 330 | U.S. | Longitudinal | | | | X | X | X |
| Wang | 2020 | Mapping out a Spectrum of the Chinese public's discrimination toward the LGBT community: results from a national survey | 3195 | China | Survey | | | X | | | |
| Weissman | 2016 | HIV Prevalence and Risk Associated with HIV Infection among Transgender Individuals in Cambodia | 891 | Cambodia | Survey | | | | | X | |
| White_Hughto | 2017 | Victimization and depressive symptomology in transgender adults: The mediating role of avoidant coping | 412 | U.S. | Survey | | | X | | | |
| Woldford-Clevenger | 2021 | Minority Stress and drug use among transgender and gender diverse adults: A daily diary study | 38 | U.S. | Longitudinal | | | | | X | |
| Wolfe | 2021 | Transgender-related discrimination and substance use, substance use disorder diagnosis and treatment history among transgender adults | 573 | U.S. | Survey | | | | | X | |

| | | | | | | | | | | |
|---|---|---|---|---|---|---|---|---|---|---|
| Wolfe | 2023 | Structural Equation Modeling of Stigma and HIV Prevention Clinical Services Among Transgender and Gender Diverse Adults: The Mediating Role of Substance Use and HIV Sexual Risk | 529 | U.S. | Survey | | X | | | |
| Yang | 2015 | Stigmatization and Mental Health in a Diverse Sample of Transgender Women | 191 | U.S. | Survey | | | | X | |
| Yang | 2016 | A cross-sectional study of associations between casual partner, friend discrimination, social support and anxiety symptoms among Chinese transgender women | 209 | China | Survey | | | | X | X |
| Yi | 2017 | HIV prevalence, risky behaviors, and discrimination experiences among transgender women in Cambodia: descriptive findings from a national integrated biological and behavioral survey | 1375 | Cambodia | Survey/Secondary Data Analysis | | | | | X |
| Yi | 2018 | Exposure to gender-based violence and depressive symptoms among transgender women in Cambodia: findings from the National Integrated Biological and Behavioral Survey 2016 | 1375 | Cambodia | Survey/Secondary Data Analysis | X | | | | X |
| Zwickl | 2021 | Factors associated with suicide attempts among Australian transgender adults | 927 | Australia | Survey | | | | | X |

### Appendix C. Risk of bias assessment

| Author, Year | Was the sample frame appropriate to address the target population? | Were study participants sampled in an appropriate way? | Was the sample size adequate? | Were the study subjects and the setting described in detail? | Was the data analysis conducted with sufficient coverage of the identified sample? | Were valid methods used for the identification of the condition? | Was the condition measured in a standard, reliable way for all participants? | Was there appropriate statistical analysis? | Was the response rate adequate, and if not, was the low response rate managed appropriately? | Overall Appraisal |
|---|---|---|---|---|---|---|---|---|---|---|
| Anderson et al. 2021 | 0 | 0 | 0 | 1 | 0 | 1 | 1 | 1 | 0 | 4 |
| Azhar et al. 2022 | 0 | 0 | 0 | 1 | 0 | 1 | 1 | 1 | 0 | 4 |
| Barr et al. 2022 | 0 | 0 | 0 | 1 | 0 | 1 | 1 | 1 | 0 | 4 |
| Basar et al. 2016a | 0 | 0 | 0 | 1 | 0 | 1 | 1 | 1 | 1 | 5 |
| Basar et al. 2016b | 0 | 0 | 0 | 1 | 0 | 1 | 1 | 1 | 1 | 5 |
| Bauermeister et al. | 0 | 0 | 0 | 1 | 0 | 1 | 1 | 1 | 0 | 4 |
| Boza et al., 2014 | 0 | 0 | 1 | 1 | 0 | 1 | 1 | 1 | 0 | 5 |
| Bretherton et al., 2021 | 0 | 0 | 1 | 1 | 0 | 1 | 1 | 1 | 0 | 5 |
| Budhwani et al., 2017_2 | 0 | 0 | 0 | 1 | 0 | 1 | 1 | 1 | 0 | 4 |
| Busby et al. | 0 | 0 | 0 | 1 | 0 | 1 | 1 | 1 | 0 | 4 |
| Caceres et al. | 0 | 0 | 0 | 1 | 0 | 1 | 1 | 1 | 0 | 4 |
| Casey et al 2019 | 0 | 1 | 0 | 1 | 0 | 1 | 1 | 1 | 0 | 5 |
| *De_Mattos_Russo_Rafael et al. 2021* | 0 | 0 | 0 | 1 | 0 | 0 | 1 | 1 | 0 | 3 |
| Fredriksen-Goldsen et al. 2013 | 0 | 0 | 0 | 1 | 0 | 1 | 1 | 1 | 0 | 4 |
| Fritz 2016 | 0 | 0 | 0 | 1 | 0 | 1 | 1 | 1 | 0 | 4 |
| Gamarel et al. 2014 | 0 | 0 | 0 | 1 | 0 | 1 | 1 | 1 | 0 | 4 |
| Gamarel et al. 2016 | 0 | 0 | 0 | 1 | 0 | 1 | 1 | 1 | 0 | 4 |
| Glick et al., 2018 | 0 | 0 | 1 | 1 | 0 | 1 | 1 | 1 | 0 | 5 |
| House et al. 2011 | 0 | 0 | 0 | 1 | 0 | 1 | 1 | 1 | 0 | 4 |
| Hsiang et al. 2022 | 0 | 0 | 1 | 1 | 0 | 1 | 1 | 1 | 0 | 4 |
| Hughto et al., 2021 | 0 | 0 | 1 | 1 | 0 | 1 | 1 | 1 | 0 | 4 |

| | | | | | | | | | | |
|---|---|---|---|---|---|---|---|---|---|---|
| Hughto et al., 2022 | 0 | 0 | 1 | 1 | 0 | 1 | 1 | 1 | 0 | 5 |
| Jaggi et al. 2018 | 0 | 0 | 1 | 0 | 0 | 1 | 1 | 1 | 0 | 5 |
| Kachen et al., 2021 | 0 | 0 | 1 | 1 | 0 | 1 | 1 | 1 | 0 | 5 |
| Kachen et al., 2022 | 0 | 0 | 1 | 1 | 0 | 1 | 1 | 1 | 0 | 4 |
| Kaplan et al., 2016 | 0 | 0 | 0 | 1 | 0 | 1 | 1 | 1 | 0 | 5 |
| Kattari et al., 2015_1 | 0 | 0 | 1 | 1 | 0 | 1 | 1 | 1 | 0 | 5 |
| Kattari et al., 2015_2 | 0 | 0 | 1 | 1 | 0 | 1 | 1 | 1 | 0 | 4 |
| Kattari et al., 2016 | 0 | 0 | 1 | 1 | 0 | 1 | 1 | 1 | 0 | 5 |
| Kattari et al., 2017 | 0 | 0 | 1 | 1 | 0 | 1 | 1 | 1 | 0 | 5 |
| Kcomt et al., 2020_2 | 0 | 0 | 1 | 1 | 0 | 1 | 1 | 1 | 0 | 5 |
| Kcomt et al., 2020_3 | 0 | 0 | 1 | 1 | 0 | 0 | 1 | 1 | 0 | 5 |
| Kidd et al. 2019 | 0 | 0 | 0 | 1 | 0 | 1 | 1 | 1 | 0 | 5 |
| Klemmer et al., 2021 | 0 | 0 | 0 | 1 | 0 | 1 | 1 | 1 | 0 | 4 |
| Lacombe-Duncan et al., 2019_1 | 0 | 0 | 0 | 1 | 0 | 1 | 1 | 1 | 0 | 4 |
| Lacombe-Duncan et al., 2019_2 | 0 | 0 | 0 | 1 | 0 | 1 | 1 | 1 | 0 | 4 |
| Lee et al., 2021 | 0 | 0 | 1 | 1 | 0 | 1 | 1 | 1 | 0 | 4 |
| Lee et al., 2022 | 0 | 0 | 0 | 1 | 0 | 1 | 1 | 1 | 0 | 4 |
| Leite et al. 2021 | 0 | 0 | 0 | 1 | 0 | 1 | 1 | 1 | 0 | 5 |
| Lerner et al., 2020 | 0 | 0 | 1 | 1 | 0 | 1 | 1 | 1 | 0 | 4 |
| Lerner et al., 2021 | 0 | 0 | 1 | 1 | 0 | 1 | 1 | 1 | 0 | 4 |
| Levine et al., 2022 | 0 | 0 | 1 | 1 | 0 | 1 | 1 | 1 | 0 | 5 |
| Lewis et al., 2019 | 0 | 0 | 1 | 1 | 0 | 1 | 1 | 1 | 0 | 5 |
| Logie et al. 2019 | 0 | 0 | 0 | 1 | 0 | 1 | 1 | 1 | 0 | 5 |
| Lozano-Verduzco et al., 2021 | 0 | 0 | 0 | 1 | 0 | 1 | 1 | 1 | 0 | 5 |
| Luz et al. 2022 | 0 | 0 | 0 | 1 | 0 | 1 | 1 | 1 | 0 | 4 |
| Marshall et al., 2016 | 0 | 0 | 1 | 1 | 0 | 1 | 1 | 1 | 0 | 4 |
| McDowell et al. 2019 | 0 | 0 | 0 | 1 | 0 | 1 | 1 | 1 | 0 | 4 |
| Messinger et al., 2022_1 | 0 | 0 | 1 | 1 | 0 | 1 | 1 | 1 | 0 | 5 |
| MezaLazaro et al., 2021 | 0 | 0 | 1 | 1 | 0 | 1 | 1 | 1 | 0 | 4 |
| Milner et al., 2019 | 0 | 0 | 1 | 1 | 0 | 1 | 1 | 1 | 0 | 5 |
| Nematollahi et al. 2022 | 0 | 0 | 0 | 1 | 0 | 1 | 1 | 1 | 0 | 5 |
| Nemoto et al., 2011 | 0 | 0 | 1 | 1 | 0 | 1 | 1 | 1 | 0 | 5 |
| Palve et al., 2018 | 0 | 0 | 1 | 1 | 0 | 1 | 1 | 1 | 0 | 4 |

| | | | | | | | | | | |
|---|---|---|---|---|---|---|---|---|---|---|
| Parr et al., 2019 | 0 | 0 | 0 | 1 | 0 | 0 | 1 | 1 | 0 | 5 |
| Poteat et al., 2017 | 0 | 0 | 1 | 1 | 0 | 1 | 1 | 1 | 0 | 5 |
| Rafael et al. 2021 | 0 | 0 | 0 | 1 | 0 | 0 | 1 | 1 | 0 | 3 |
| Ralston et al. 2022 | 0 | 0 | 0 | 0 | 0 | 1 | 1 | 1 | 0 | 5 |
| Rodriguez-Madera et al., 2017 | 0 | 0 | 0 | 1 | 0 | 1 | 1 | 1 | 0 | 3 |
| Romanelli et al., 2018 | 0 | 0 | 1 | 1 | 0 | 1 | 1 | 1 | 0 | 3 |
| Romanelli et al., 2020 | 0 | 0 | 1 | 1 | 0 | 1 | 1 | 1 | 0 | 4 |
| Rood et al. 2015 | 0 | 0 | 0 | 1 | 0 | 0 | 1 | 1 | 0 | 5 |
| Rosentel et al., 2021 | 0 | 0 | 1 | 1 | 0 | 1 | 1 | 1 | 0 | 5 |
| Rotondi et al. 2011 | 0 | 0 | 0 | 1 | 0 | 1 | 1 | 1 | 0 | 3 |
| Rouhani et al. 2021 | 0 | 0 | 0 | 1 | 0 | 1 | 1 | 1 | 0 | 5 |
| Ruggs et al. 2015 | 0 | 0 | 0 | 0 | 0 | 0 | 1 | 1 | 0 | 4 |
| Salas-Espinoza et al., 2017 | 0 | 0 | 1 | 1 | 0 | 1 | 1 | 1 | 0 | 4 |
| Salazar et al. 2017 | 0 | 0 | 0 | 1 | 0 | 0 | 1 | 1 | 0 | 2 |
| Scandurra et al. 2020 | 0 | 0 | 0 | 1 | 0 | 1 | 1 | 1 | 0 | 5 |
| Scheim et al. 2017_b | 0 | 0 | 0 | 1 | 0 | 1 | 1 | 1 | 0 | 3 |
| Scheim et al., 2016 | 0 | 0 | 1 | 1 | 0 | 1 | 1 | 1 | 0 | 4 |
| Schweizer et al. 2022 | 0 | 0 | 0 | 1 | 0 | 0 | 1 | 1 | 0 | 4 |
| Seelman et al. 2016 | 0 | 0 | 0 | 1 | 0 | 1 | 1 | 1 | 0 | 5 |
| Seelman et al., 2020 | 0 | 0 | 1 | 1 | 0 | 1 | 1 | 1 | 0 | 3 |
| Sevelius et al. 2014 | 0 | 0 | 0 | 1 | 0 | 0 | 1 | 1 | 0 | 4 |
| Shah et al., 2018 | 0 | 0 | 1 | 1 | 0 | 1 | 1 | 1 | 0 | 5 |
| She et al. 2021_1 | 0 | 0 | 0 | 1 | 0 | 1 | 1 | 1 | 0 | 3 |
| She et al. 2021-2 | 0 | 0 | 0 | 1 | 0 | 1 | 1 | 1 | 0 | 5 |
| Shires et al., 2015 | 0 | 0 | 0 | 1 | 0 | 1 | 1 | 1 | 0 | 4 |
| Siamisang et al., 2022 | 0 | 0 | 1 | 1 | 0 | 1 | 1 | 1 | 0 | 4 |
| Silva et al., 2021 | 0 | 0 | 1 | 1 | 0 | 1 | 1 | 1 | 0 | 4 |
| Silva et al., 2022 | 0 | 0 | 0 | 1 | 0 | 1 | 1 | 1 | 0 | 5 |
| Sutherland et al., 2021 | 0 | 0 | 1 | 1 | 0 | 1 | 1 | 1 | 0 | 5 |
| Tabaac et al. 2018 | 0 | 0 | 0 | 1 | 0 | 1 | 1 | 1 | 0 | 4 |
| Thompson-Blum et al. | 0 | 0 | 0 | 1 | 0 | 1 | 1 | 1 | 0 | 5 |
| Valente et al. 2022 | 0 | 0 | 0 | 1 | 0 | 0 | 1 | 1 | 0 | 4 |
| Wang et al. 2020 | 0 | 0 | 1 | 1 | 0 | 0 | 1 | 1 | 0 | 4 |
| Weissman et al. 2016 | 0 | 0 | 1 | 1 | 0 | 0 | 1 | 1 | 0 | 3 |

| | | | | | | | | | | |
|---|---|---|---|---|---|---|---|---|---|---|
| White_Hughto et al. 2017 | 0 | 0 | 0 | 1 | 0 | 1 | 1 | 1 | 0 | 4 |
| Woldford-Clevenger et al. 2021 | 0 | 0 | 0 | 1 | 0 | 1 | 1 | 1 | 0 | 4 |
| Wolfe et al. 2021 | 0 | 0 | 1 | 1 | 0 | 1 | 1 | 1 | 0 | 4 |
| Wolfe et al. 2023 | 0 | 0 | 1 | 1 | 0 | 1 | 1 | 1 | 0 | 4 |
| Yang et al. 2015_2 | 0 | 0 | 0 | 1 | 0 | 1 | 1 | 1 | 0 | 5 |
| Yang et al., 2016_1 | 0 | 0 | 1 | 1 | 0 | 1 | 1 | 1 | 1 | 5 |
| Yi et al., 2017 | 0 | 0 | 1 | 1 | 0 | 1 | 1 | 1 | 0 | 4 |
| Yi et al., 2018 | 0 | 0 | 1 | 1 | 0 | 1 | 1 | 1 | 0 | 6 |
| Zwickl et al., 2021 | 0 | 0 | 1 | 1 | 0 | 1 | 1 | 1 | 0 | 5 |

**Appendix D. Forest plots showing prevalence of discrimination across different sectors experienced by trans and gender-diverse adults**

D1. Discrimination in Housing

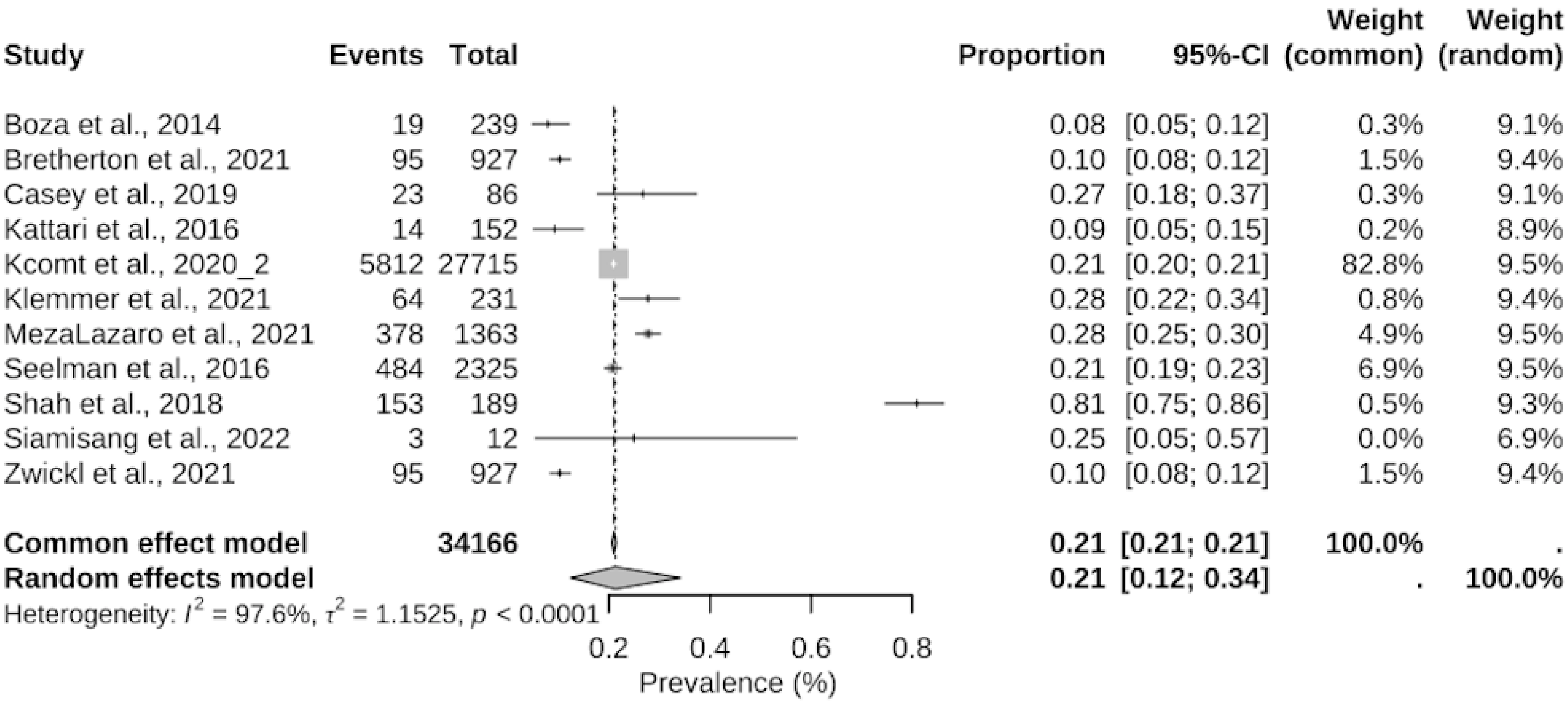

D2. Discrimination in Healthcare

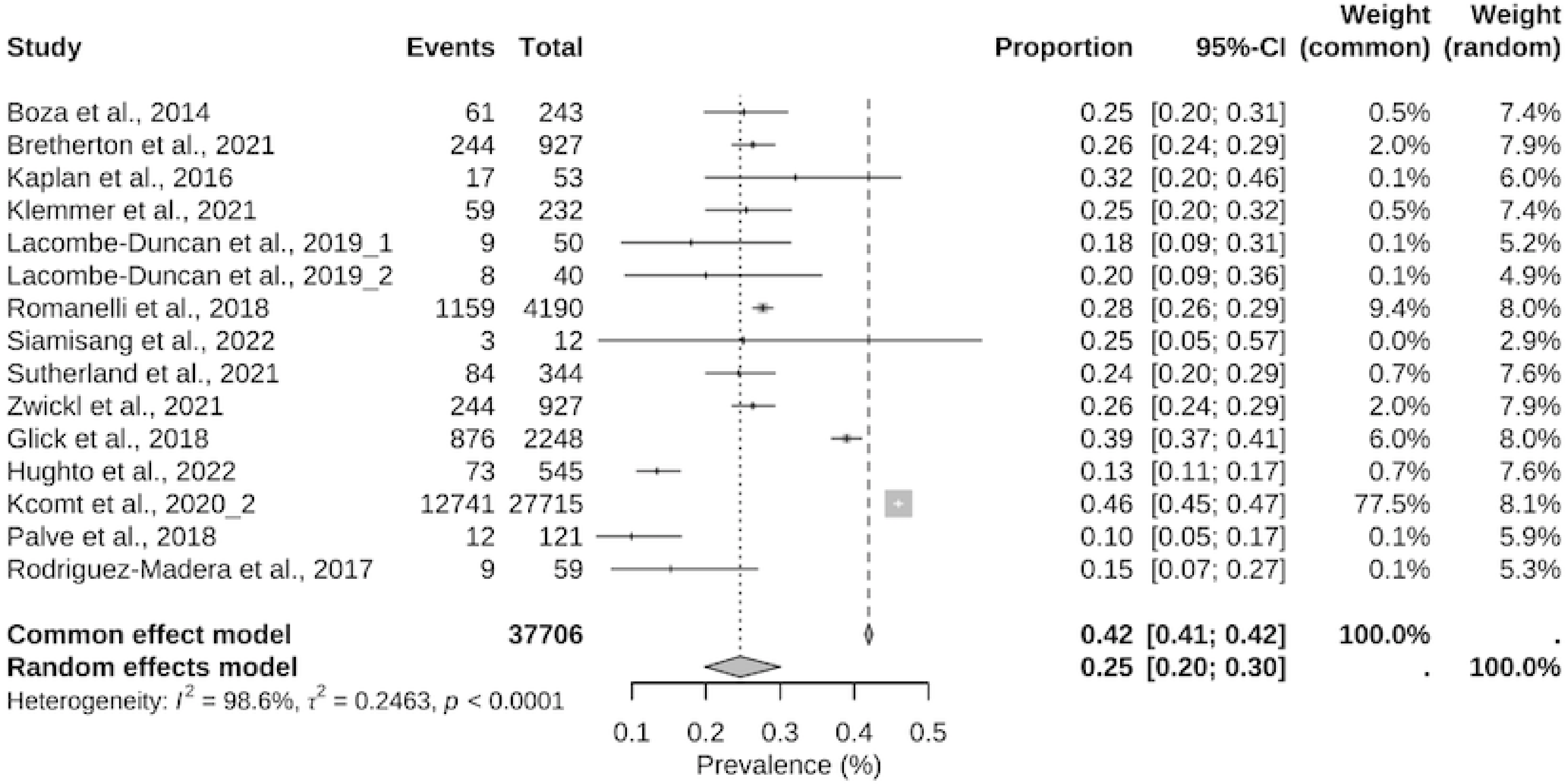

D3. Discrimination in Employment

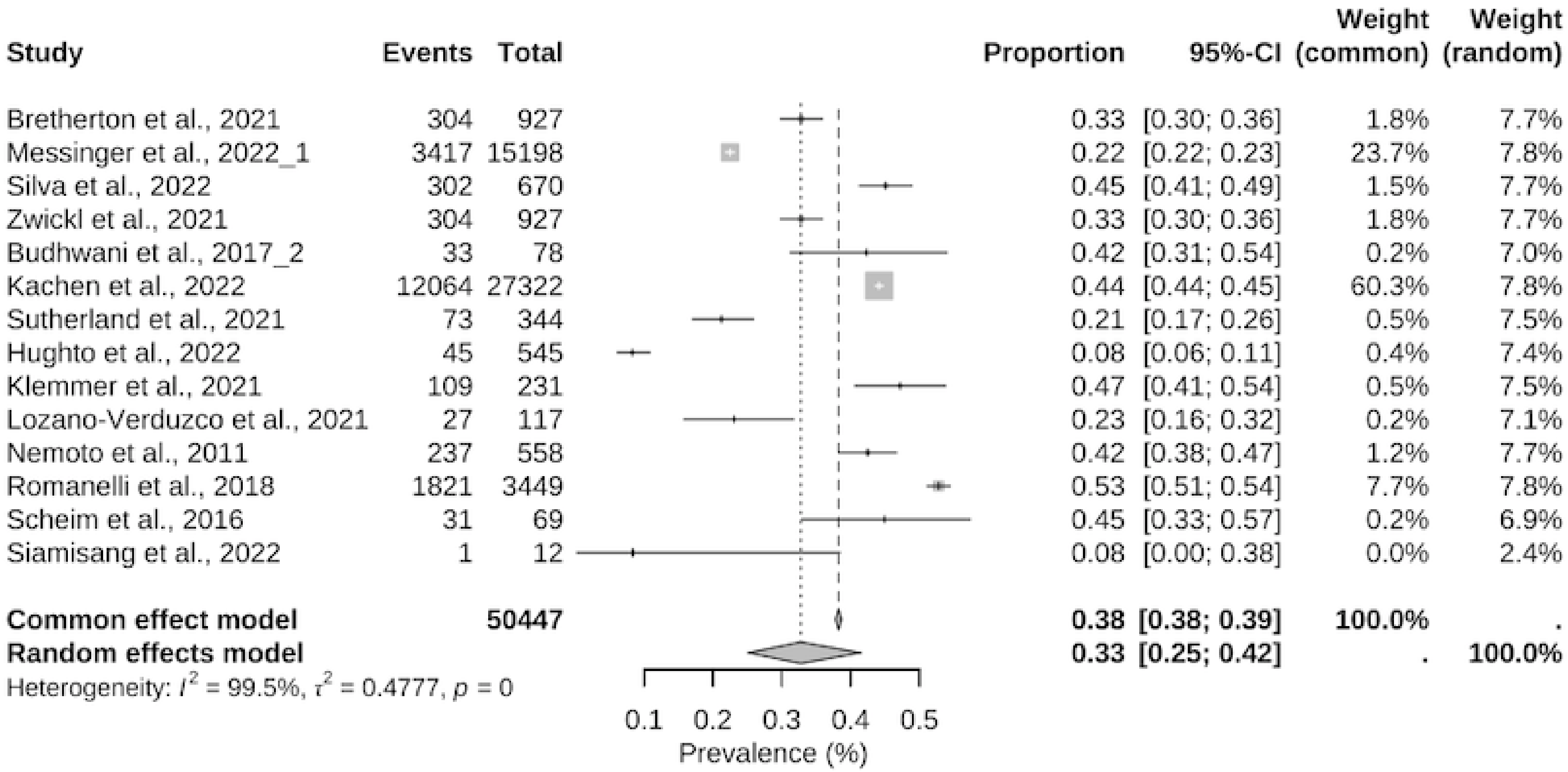

D4. Discrimination in Employment: subgroup: Fired/Denied Job/Denied Promotion

| Study | Events | Total | Proportion | 95%-CI | Weight (common) | Weight (random) |
|---|---|---|---|---|---|---|
| Hughto et al., 2022 | 45 | 545 | 0.08 | [0.06; 0.11] | 3.6% | 15.5% |
| Klemmer et al., 2021 | 109 | 231 | 0.47 | [0.41; 0.54] | 5.1% | 15.6% |
| Lozano-Verduzco et al., 2021 | 27 | 117 | 0.23 | [0.16; 0.32] | 1.8% | 15.1% |
| Nemoto et al., 2011 | 237 | 558 | 0.42 | [0.38; 0.47] | 12.0% | 15.8% |
| Romanelli et al., 2018 | 1821 | 3449 | 0.53 | [0.51; 0.54] | 75.8% | 15.9% |
| Scheim et al., 2016 | 31 | 69 | 0.45 | [0.33; 0.57] | 1.5% | 14.9% |
| Siamisang et al., 2022 | 1 | 12 | 0.08 | [0.00; 0.38] | 0.1% | 7.3% |
| **Common effect model** | | **4981** | **0.48** | **[0.47; 0.50]** | **100.0%** | **.** |
| **Random effects model** | | | **0.31** | **[0.17; 0.49]** | **.** | **100.0%** |

Heterogeneity: $I^2 = 98.0\%$, $\tau^2 = 0.9310$, $p < 0.0001$

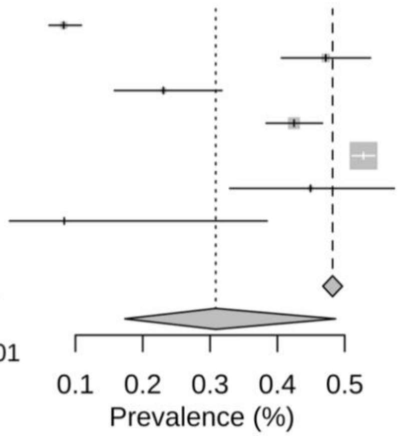

D5. Discrimination in Interactions with Police/Prison System

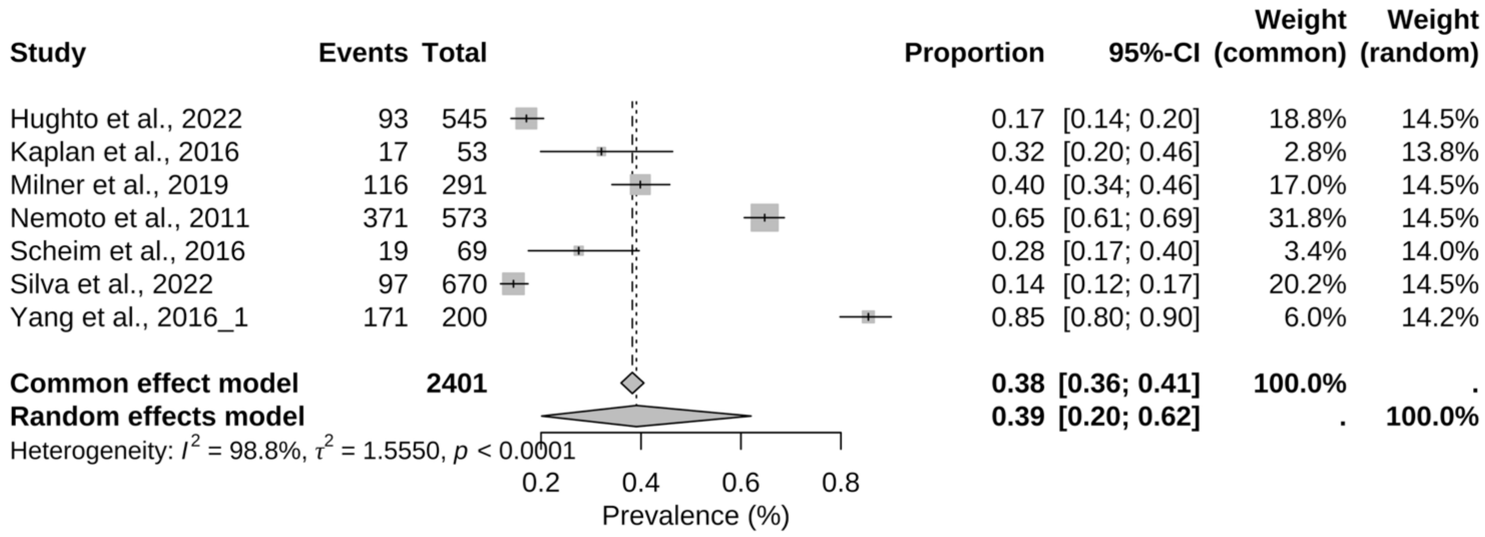
